# Visceral Leishmaniasis in pregnancy and vertical transmission: A systematic literature review on the therapeutic orphans

**DOI:** 10.1101/2021.04.16.21255552

**Authors:** Prabin Dahal, Sauman Singh-Phulgenda, Brittany J Maguire, Eli Harriss, Koert Ritmeijer, Fabiana Alves, Philippe J Guerin, Piero L Olliaro

**Author notes:** Correspondence to Infectious Diseases Data Observatory (IDDO), Oxford, UK. Email addresses: PD, SSP, BJM, EH, KR, FA, PJG, PLO.

## Abstract

**Background:** Reports on the occurrence and outcome of Visceral Leishmaniasis (VL) in pregnant women is rare in published literature. The occurrence of VL in pregnancy is not systematically captured and cases are rarely followed-up to detect consequences of infection and treatment on the mother and foetus.

**Methods:** A review of all published literature was undertaken to identify cases of VL infections during pregnancy by searching the following database: Ovid MEDLINE®; Ovid Embase; Cochrane Database of Systematic Reviews; Cochrane Central Register of Controlled Trials; World Health Organization Global Index Medicus: LILACS (Americas); IMSEAR (South- East Asia); IMEMR (Eastern Mediterranean); WPRIM (Western Pacific); ClinicalTrials.gov; and the WHO International Clinical Trials Registry Platform. Selection criteria included any clinical reports describing the disease in pregnancy or vertical transmission of the disease in humans. Articles meeting pre-specified inclusion criteria and non-primary research articles such as textbook, chapters, letters, retrospective case description, or reports of accidental inclusion in trials were also considered.

**Results:** We screened 272 publications and identified a total of 70 records (1926–2020) describing 447 VL cases in pregnant women. The disease was detected during pregnancy in 394 (88.1%), retrospectively confirmed after giving birth in 52 (11.6%), and the time of identification was not clear in 1 (0.2%). Of the 394 mothers whose infection was identified during pregnancy, 344 (89.1%) received a treatment, 3 (0.8%) were untreated, and the treatment status was not clear in the remaining 47 (12.2%). Of 344 mothers, Liposomal Amphotericin B (L-AmB) was administered in 202 (58.7%) and pentavalent antimony (PA) in 92 (26.7%). Outcomes were reported in 176 mothers treated L-AmB with 4 (2.3%) reports of maternal deaths, 5 (2.8%) miscarriages, and 2 (1.1%) foetal death/stillbirth. For PA, outcomes were reported in 87 mothers of whom 4 (4.6%) died, 24 (27.6%) had spontaneous abortion, 2 (2.3%) had miscarriages. A total of 26 cases of confirmed, probable or suspected cases of vertical transmission were identified and the median time to detection was 6 months (range: 0–18 months).

**Conclusions:** Outcomes of VL treatment during pregnancy is rarely reported and under- researched. When it is reported, information is often incomplete and it is difficult to derive generalisable information on outcomes for mothers and babies, although reported data favours the usage of liposomal amphotericin B for the treatment of VL in pregnant women.

**Author summary:** Visceral Leishmaniasis (VL) is a neglected tropical disease with an estimated incidence of 50,000 to 90,000 cases in 2019. Women who are susceptible to becoming pregnant or those who are pregnant and lactating are regularly excluded from clinical studies of VL. A specific concern of public health relevance is the little knowledge of the consequences of VL and its treatment on the mother and the foetus. We did a systematic review of all published literature with an overarching aim of identifying cases of VL in pregnancy and assess the risk-benefit balance of antileishmanial therapies to the mother and the child. We identified a total of 70 records (1926–2020) describing 447 VL cases in pregnant women. In 394 mothers, infection was identified during pregnancy of whom 202 received Liposomal Amphotericin B (L-AmB) and 92 received pentavalent antimony (PA). Reports of maternal deaths, abortion, and miscarriages were proportionally lower among those who received L- AmB compared to PA regimens. A total of 26 cases of confirmed, probable or suspected cases of vertical transmission were identified and the median time to detection was 6 months (range: 0–18 months). Our review brings together scattered observations of VL in pregnant women in the clinical literature and clearly highlights that the disease in pregnancy is under-reported and under-studied. Our findings indicate that L-AmB should be the preferred treatment for VL during pregnancy.

## Introduction

Visceral Leishmaniasis (VL) is a neglected tropical disease caused by Leishmania sp. parasites transmitted by female sandflies. The disease is endemic in parts of South Asia, East Africa, South America and the Mediterranean region with an estimated 50,000 to 90,000 cases in 2019 [1]. A specific concern of public health relevance is the little knowledge of the clinical aspects of VL and treatment outcomes in pregnant and lactating women [2].

In pregnancy, VL diagnosis relies essentially on symptoms and serology as parasite detection by splenic aspiration is not recommended because of the risk for the foetus. More severe anaemia and increased requirements for blood transfusion have been reported for pregnant women infected with VL [2]. Case management must take into account the consequences of the disease and the therapeutic intervention on the mother-foetus pair [3]. Of note, except amphotericin B, all other available drugs are either contraindicated or subjected to restricted use in pregnant and lactating women and in women of child-bearing age (Table 1) [4–6]. Further complexities arise from potential vertical transmission of the disease either congenitally (maternal–foetal transmission in utero) or through transplacental infection as a result of blood exchange during labour. While vertical transmission of VL is well-studied and established in animal studies, reports in humans are sporadic with observations of clinical manifestation several months post-partum [7–10]. Such vertical transmission can induce in utero death or can be potentially deleterious to the foetus and infant [6,9,11].

**Table 1.** Antileishmanial usage during pregnancy

| Drug | Indication | FDA Category<br>(reviewed in [23,24]) |
| --- | --- | --- |
| Pentavalent antimonials:<br>Pentostam<br>(Sodium Stibogluconate) | “Although no effects on the foetus have been reported, Pentostam should be withheld during pregnancy unless the potential benefits to the patient outweigh the possible risk to the foetus. Children should not be breast-fed by mothers receiving Pentostam”<br>– Source: The EMC [25]<br>“Pentavalent antimonials are less safe in pregnancy, as they can result in spontaneous abortion, preterm deliveries and hepatic encephalopathy in the mother and vertical transmission”<br>– Source: WHO-2010 [26] | **C**<br>(Risks cannot be ruled out) |
| Amphotericin B deoxycholate | “Animal reproduction studies have failed to demonstrate a risk to the foetus and there are no adequate and well-controlled studies in pregnant women.”<br>“Amphotericin B deoxycholate and lipid formulations are the best therapeutic options for visceral leishmaniasis. No abortions or vertical transmission have been reported in mothers treated with liposomal amphotericin”<br>– Source: WHO-2010 [26] | **B**<br>(No evidence of risk in studies) |
| Liposomal amphotericin B<br>(AmBisome) | “Animal studies do not indicate direct or indirect harmful effects with respect to reproductive toxicity. The safety of AmBisome in pregnant women has not been established. Systemic fungal infections have been successfully treated in pregnant women with conventional amphotericin B without obvious effect on the foetus, but the number of cases reported is insufficient to draw any conclusions on the safety of AmBisome in pregnancy. AmBisome should only be used during pregnancy if the possible benefits to be derived outweigh the potential risks to the mother and foetus. It is unknown whether AmBisome is excreted in human breast milk. A decision on whether to breastfeed while receiving AmBisome should take into account the potential risk to the child as well as the benefit of breast feeding for the child and the benefit of AmBisome therapy for the mother”<br>–Source: The EMC [27]<br>“Amphotericin B deoxycholate and lipid formulations are the best therapeutic options for visceral leishmaniasis. No abortions or vertical transmission have been reported in mothers treated with liposomal amphotericin”<br>– Source: The WHO-2010 [26]<br>This is the first line therapy for treatment against pregnancy in Kenya, Ethiopia, Somalia, Sudan, South Sudan, Uganda, and Brazil<br>– Source: The WHO [28] | **B**<br>(No evidence of risk in studies) |
| Pentamidine | Contraindicated during the first trimester of pregnancy<br>– Source: WHO-2010 [26] | **C**<br>(Risks cannot be ruled out) |
| Miltefosine (Impavido) | Contraindicated in pregnancy: "Impavido may cause foetal harm. Foetal death and teratogenicity occurred in animals administered miltefosine at doses lower than the recommended human dose. Do not administer IMPAVIDO to pregnant women. Obtain a serum or urine pregnancy test in females of reproductive potential prior to prescribing IMPAVIDO. Females of reproductive potential should be advised to use effective contraception during IMPAVIDO therapy and for 5 months after therapy"<br>– Source: The US FDA Impavido prescribing information [29]<br>"Miltefosine is potentially embryotoxic and teratogenic and should not be used during pregnancy. Women of child-bearing age should be tested for pregnancy before treatment and use effective contraception for 3 months after treatment"<br>– Source: WHO-2010 [26] | **D**<br>(Positive evidence or risk) |
| Paromomycin (aminosidine) | "Ototoxicity in the foetus is the main concern. Insufficient data are available on the use of paromomycin in pregnant women"<br>– Source: WHO-2010 [26]<br>"Paromomycin crosses the placenta and can cause renal and auditory damage in the unborn child. Paromomycin is excreted in breast milk and adverse effects in the breastfed infant cannot be excluded."<br>– Source: National guidelines of Kenya-2017 [30] | **No category assigned** |

The regulatory restrictions and limited evidence on safety of antileishmanial chemotherapeutics on the mother-foetus pair meant that historically clinicians had to rely on personal experience or limited published case-reports to make a decision. This led to some clinicians delaying the treatment of pregnant women until after delivery, especially when the case was detected closer to the due date [12, 13]. Others had treated them when the adjudicated risk of VL to the mother outweighed the risk posed by the drug to the mother-foetus pair [14]. Similar delays in treatment of pregnant mothers has also been reported in post kala-azar dermal leishmaniasis (PKDL) [15, 16]. Currently liposomal amphotericin B (L-AmB) remains the preferred regimen for the treatment in pregnancy (Table 1). However, pregnant and lactating women are regularly excluded from clinical studies [17] and are considered “therapeutic orphans” [18]. In studies that enrol females of childbearing age, counselling measures are usually set in place to inform the patients regarding the potential teratogenic harms of study drugs and either adoption of suitable contraception methods or observance of abstinence is mandatory (for example in miltefosine trials) [17]. In regular clinical practice and non-clinical trial settings, pregnancy tests and counselling however, might not be done routinely. A study conducted in South Asia found that only one in every six doctors ruled out pregnancy before prescribing miltefosine [19].

Finally, there is a lack of active pregnancy registries for most of the antileishmanial drugs expect for miltefosine. In the context of Impavo® (Profounda Inc.), the commercial name of miltefosine registered to the US medicines regulatory agency (US Food Drug Administration), a pregnancy registry was established to fulfil post marketing requirements [20, 21]. The recruitment of pregnant women as part of the observational study started in 2015 and the study is expected to be completed in 2026, and is estimated to recruit 0–1 patients per year over the 10 year study period, hence unlikely to generate a large volume of new safety data [20]. There are no other active pregnancy registries on exposures to VL treatments from which to derive information on consequences on gestation, mother, foetus, and the newborn. Therefore, to understand the risks and benefits of treatment to the mother and the child, one must turn to the published literature.

The most comprehensive reviews on VL in pregnant women were conducted in the mid 2000s [9, 22]. We therefore conducted a systematic review of all published literature with an overarching aim of identifying cases of VL in pregnancy. The specific objectives were to assess the risk-benefit balance of antileishmanial therapies to the mother and the child and to identify the cases of vertical transmission. The review was not limited by language or any interventions.

## Material and Methods

### Literature search

A review of all published literature was undertaken on 26th of March 2020 to identify records describing VL in pregnant women or any reports of vertical transmission of the disease in humans by searching the following clinical databases: Ovid MEDLINE®; Ovid Embase; Cochrane Database of Systematic Reviews; Cochrane Central Register of Controlled Trials; World Health Organization Global Index Medicus: LILACS (Americas); IMSEAR (South- East Asia); IMEMR (Eastern Mediterranean); WPRIM (Western Pacific); ClinicalTrials.gov; and the WHO International Clinical Trials Registry Platform (ICTRP). The systematic review was conducted in accordance with the Preferred Reporting Items for Systematic-Reviews and Meta-Analyses (PRISMA) guidelines (S1 Text)[31]. In addition, full text screening of the publications indexed in the Infectious Diseases Data Observatory (IDDO) clinical trials library was carried out to identify any description of VL in pregnant women [32]. The references of all included publications were further checked to identify any relevant articles. This review is not registered and the protocol describing the search strategy including database search strings, search dates and eligibility criteria for screening is presented in supplemental file (S2 Text).

### Study screening

Study screening was carried out in two stages to identify the studies fulfilling the inclusion and exclusion criteria (S2 Text): title and abstract screening (stage I) and then full-text screening (stage II). As reports on VL in pregnancy are sparse, articles meeting minimal inclusion criteria and non-primary research articles such as opinion pieces, clinical guidelines, textbooks, chapters, correspondences, reports of accidental inclusion in trials, or case reports of unplanned pregnancies during the study follow-up were also considered for comprehensiveness. No restrictions were applied regarding study design, follow-up duration, sample size, region, or the treatment regimen for eligibility of inclusion in this review. Title and abstracts were screened in the first stage, followed by screening the full-texts. Articles that were not in English language (Spanish, Portuguese, Korean, and German) were evaluated using google translation (https://translate.google.co.uk/).

The articles were screened against eligibility criteria by a single reviewer (PD). A second reviewer was consulted (SSP) when the first reviewer couldn’t reliably assess the eligibility. The first reviewer (PD) extracted data from all the eligible records and it was verified by the second reviewer (SSP) (who was not blinded) on all publications included in the review. Any discrepancy in the extracted information was flagged by the second reviewer and the differences were resolved through consensus. Screening and data extraction was carried out on a prospectively designed Excel database.

### Data extraction

The following bibliographic information were extracted: study title, name of the first author, year of publication, name of the study site and country. The following maternal and child characteristics were extracted: age of the mother, period of gestation (or trimester), treatment administered including drug dosage, follow-up duration, the outcome of the treatment for mother (cured, relapsed, death), and foetal outcomes (abortion, stillbirth, premature birth, healthy born, vertical transmission).

### Definitions

The records were classified as: case report/case series, prospective cohort or retrospective cohort studies. Records describing one or a small group of patients included as a part of prospective (or retrospective) studies in which VL in pregnancy was not of primary focus were considered as case report/case series. Similarly, studies that described a cohort of pregnant women without selection of a non-pregnant comparator group were also considered as case series. Countries were classified into sub-regions according to United Nations designation of geographical regions [33].

### Data analysis

Since majority of the studies included were either case reports or case series, analysis of data was restricted to presentation of descriptive statistics and meta-analysis was not carried out. Descriptive summaries were presented for the characteristics of the studies included in the review, maternal characteristics (trimester, gestational age), treatment regimen including dosage and duration, clinical outcomes on the mother and the child. Graphics were generated using R software [34].

### Assessment of risk of bias

The risk of bias in case report/case series was assessed using a checklist proposed in Murad- 2018 [35]. The following domain were assessed: patient selection, ascertainment of exposure, outcome assessment, adequacy of follow-up, and reporting of results. A single case report was considered to be at a high risk of selection bias whereas a series of cases selected based on an audit of complete records over a study period was considered to be at a low risk of selection bias. Bias in ascertainment of exposure was considered to be high if the diagnosis of VL was based solely on clinical features. For cohort studies (prospective or retrospective), risk of bias was assessed using The Newcastle-Ottawa scale. Two authors (PD, SSP) independently assessed the risk of bias in the studies included.

## Results

We identified 395 records from the literature searches up until 26th of March 2020, of which 272 were unique after removing duplicate entries. Of the 272 unique records, 99 were excluded at title and abstract screening stage (Fig 1) leaving 173 records for full-text assessment of which 53 met the eligibility criteria for inclusion in the review (Fig 1). An additional 17 records were identified by searching the references of the eligible records and through personal communication. A total of 70 records published from 1926 through 2020 were included in this review of which were 69 were case-reports or case-series and 1 was a retrospective cohort study with non-pregnant patient as a comparative group (Table 2). Further details on the studies included in this review are presented in supplemental files (S1 Data, S2 Data).

**Fig 1:**
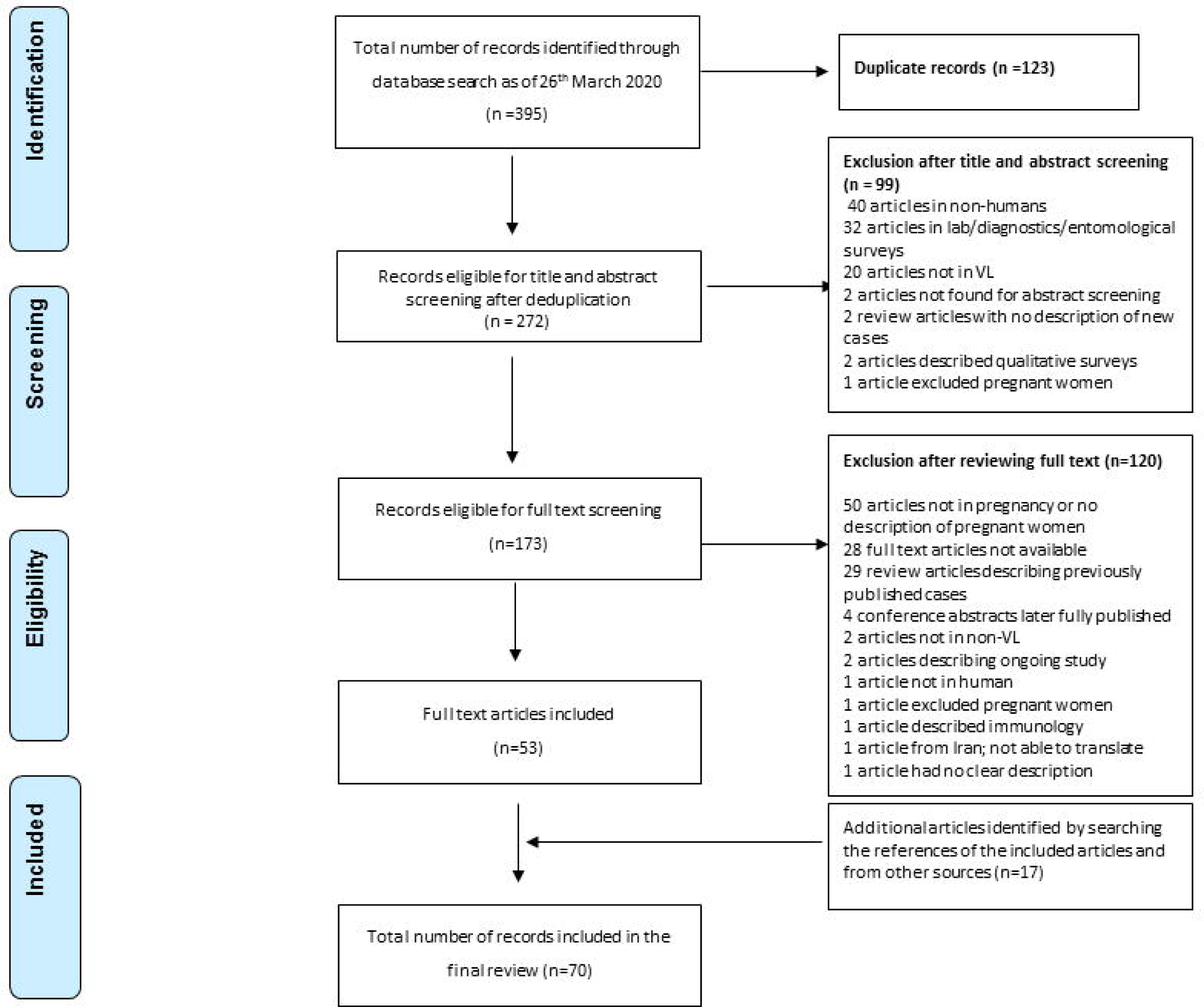
Preferred Reporting Items for Systematic Reviews and Meta-Analyses (PRISMA) flow diagram of publications screened

**Table 2:**
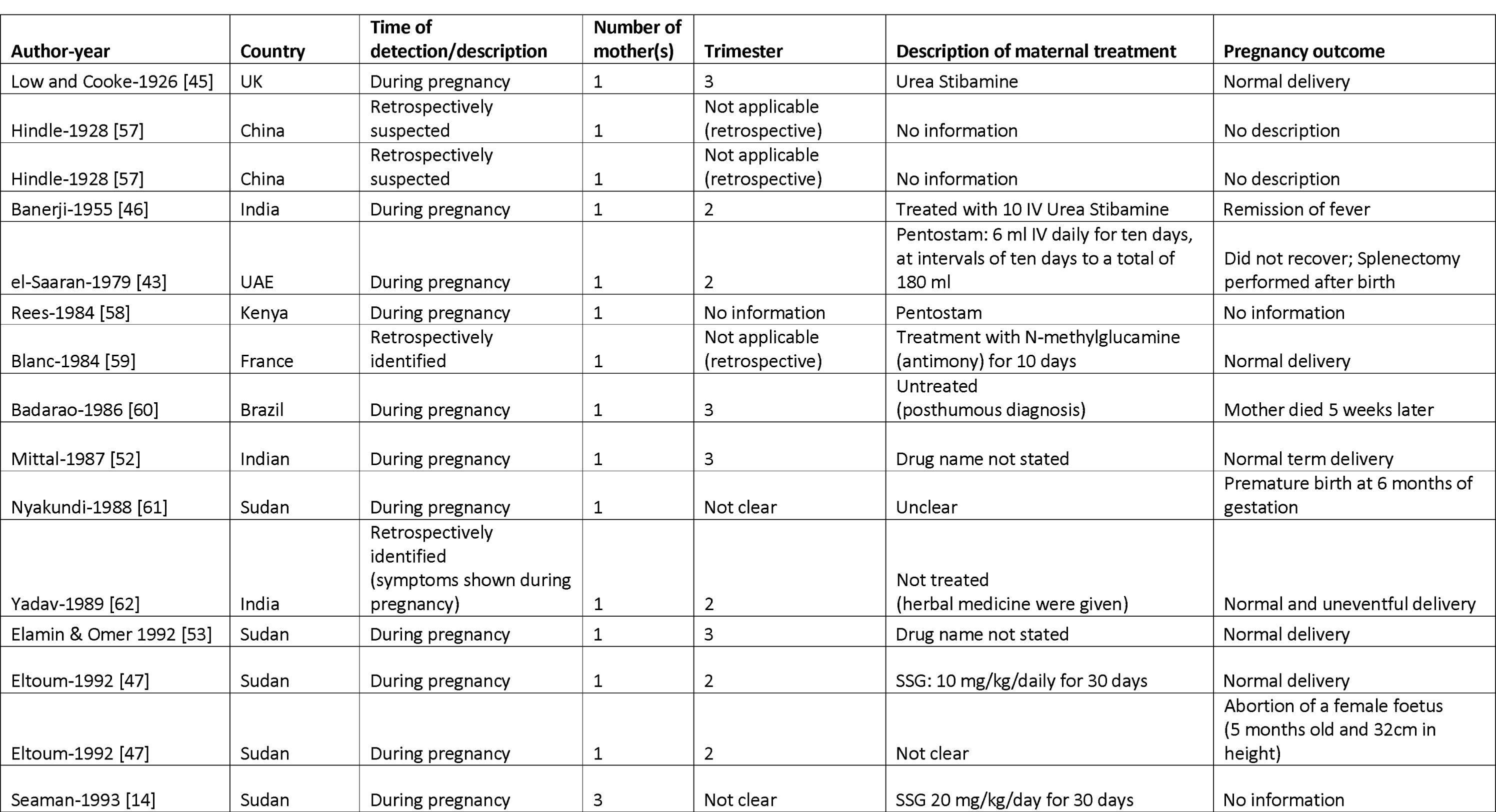
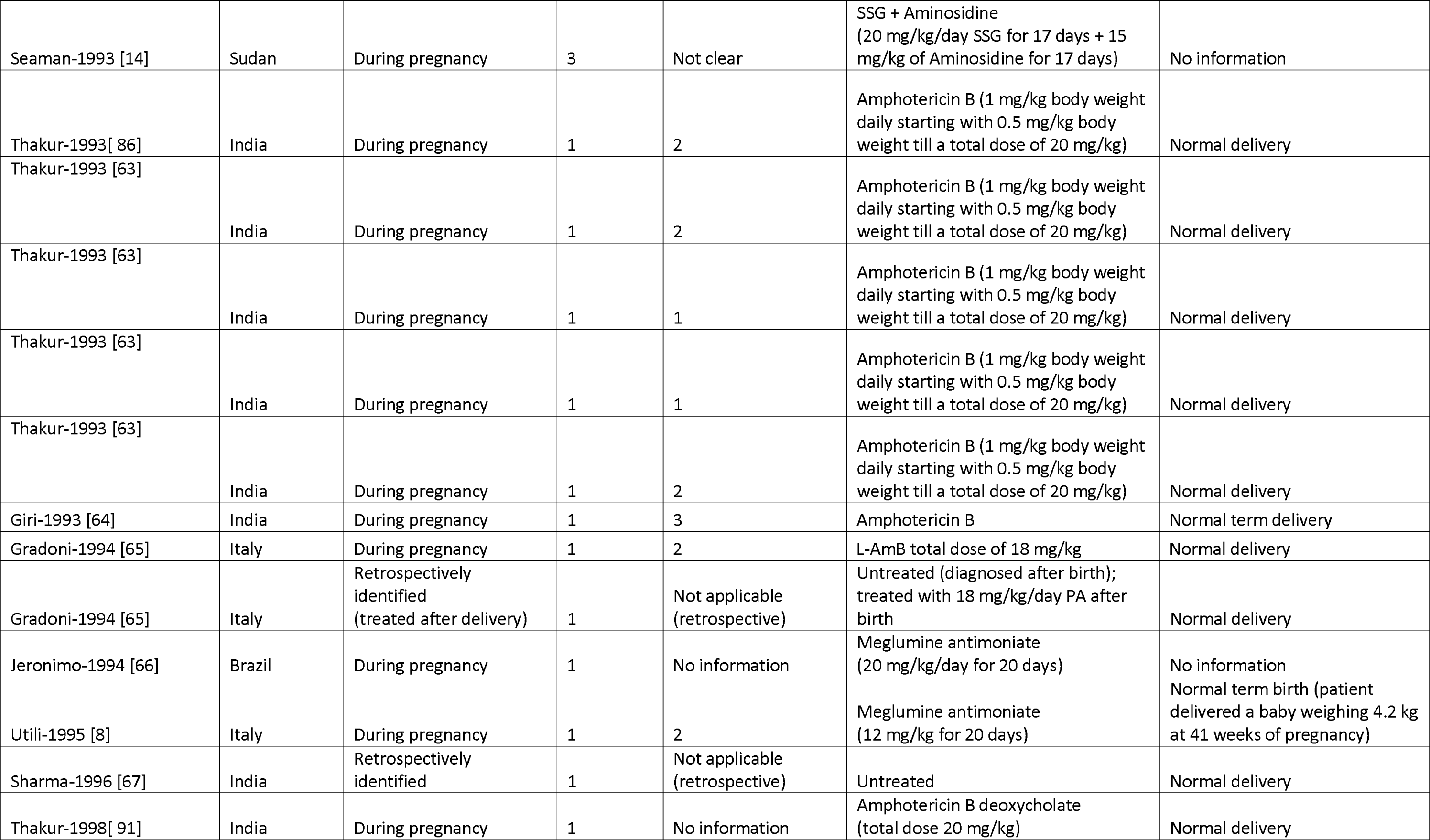
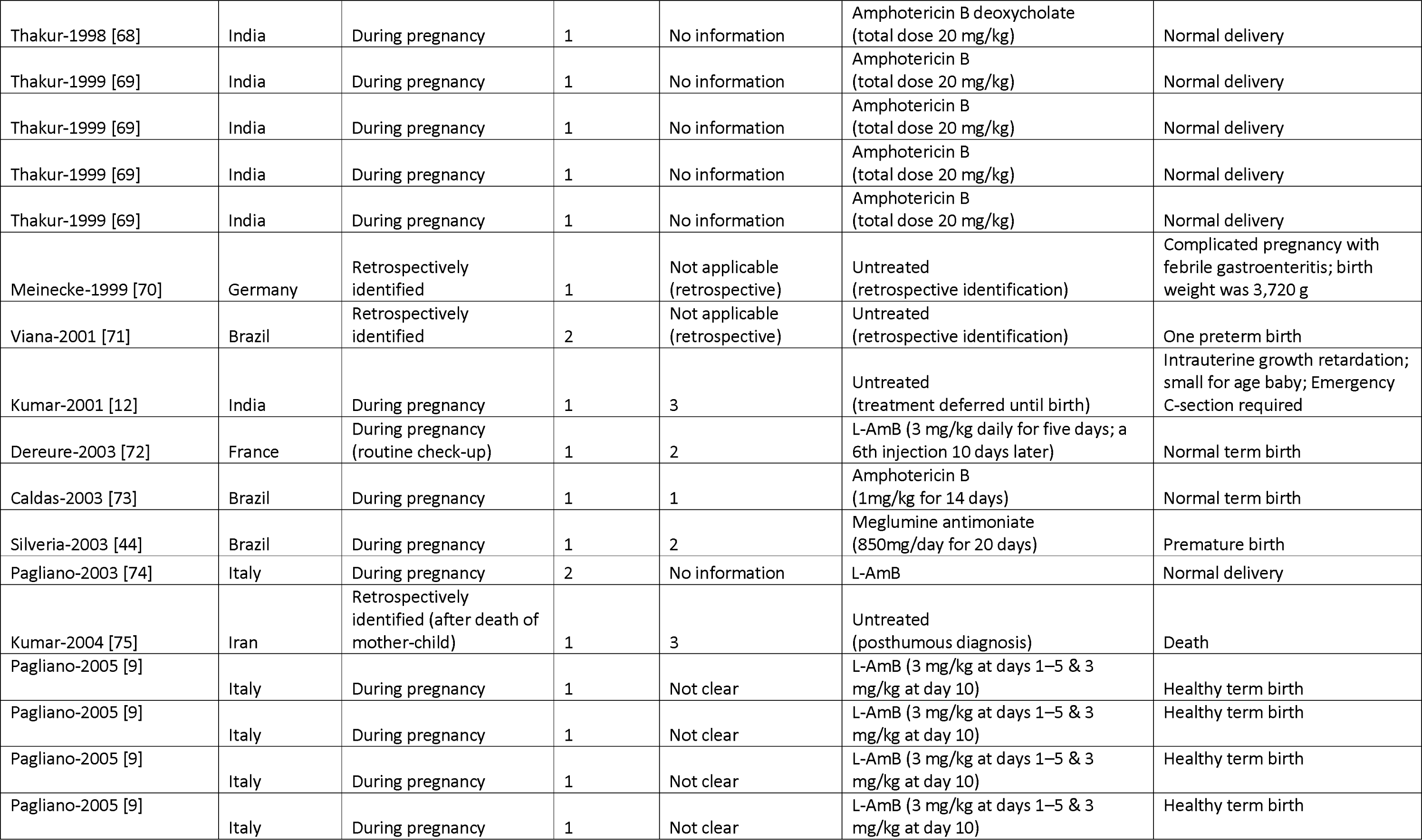
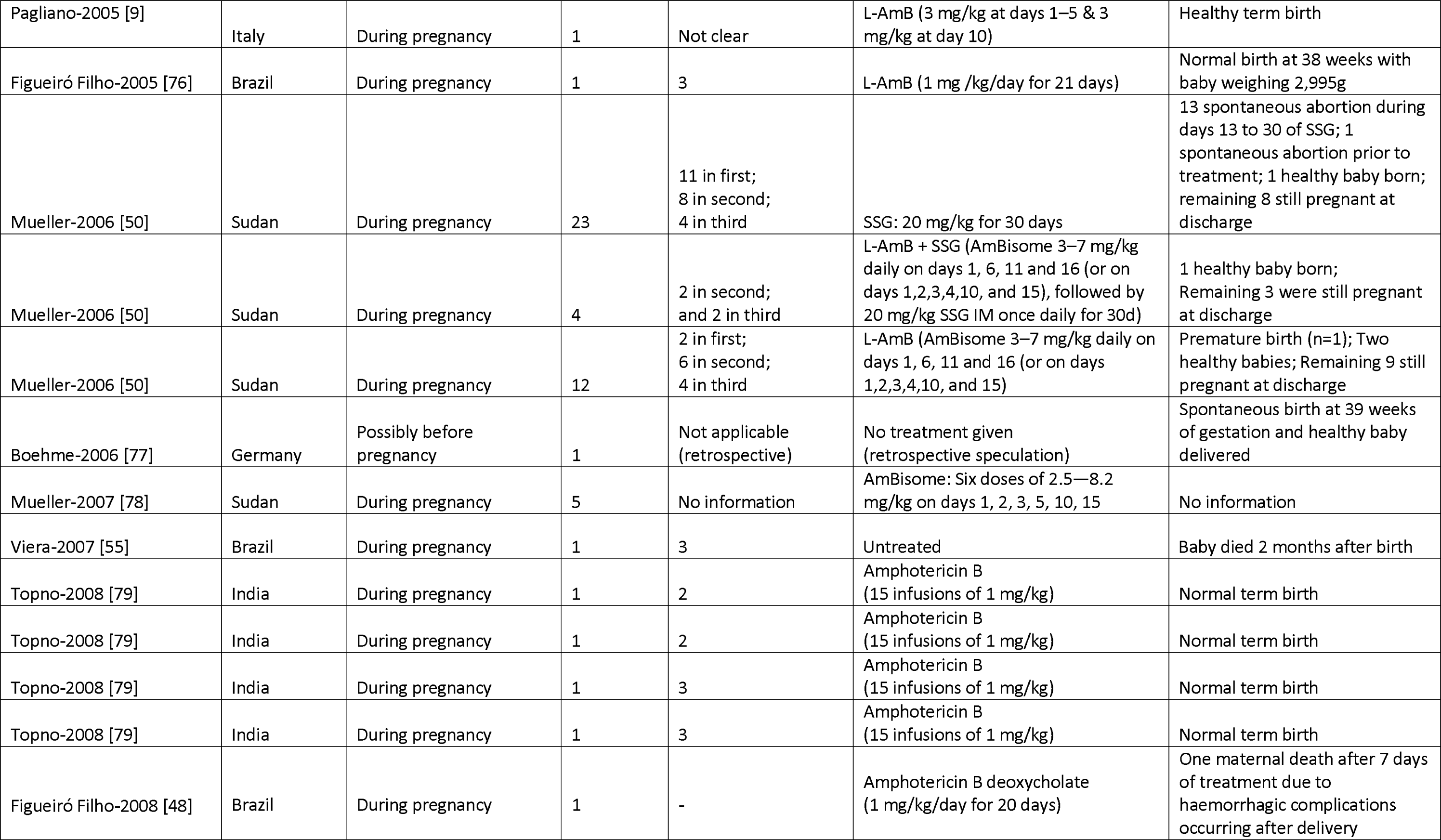
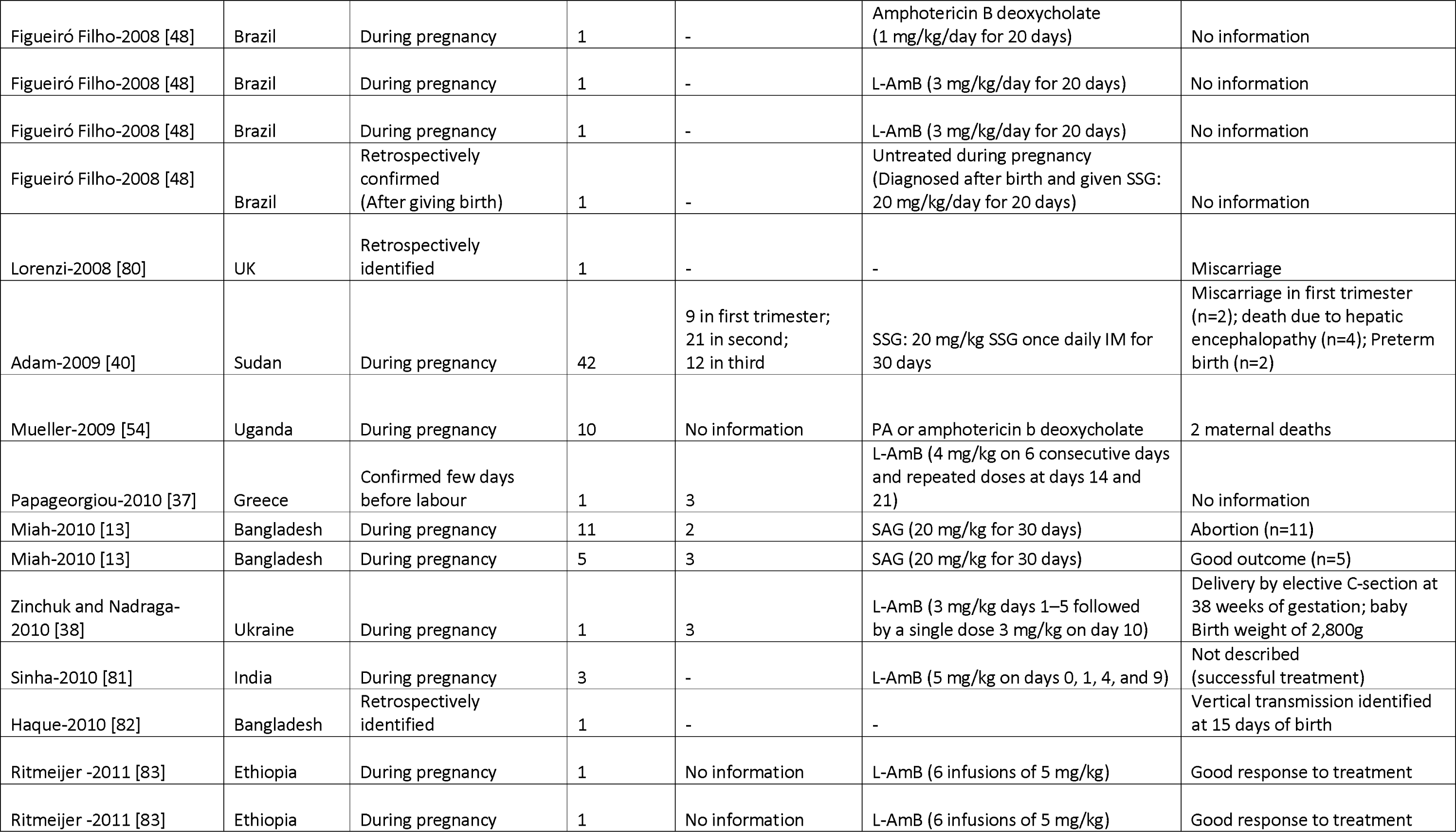
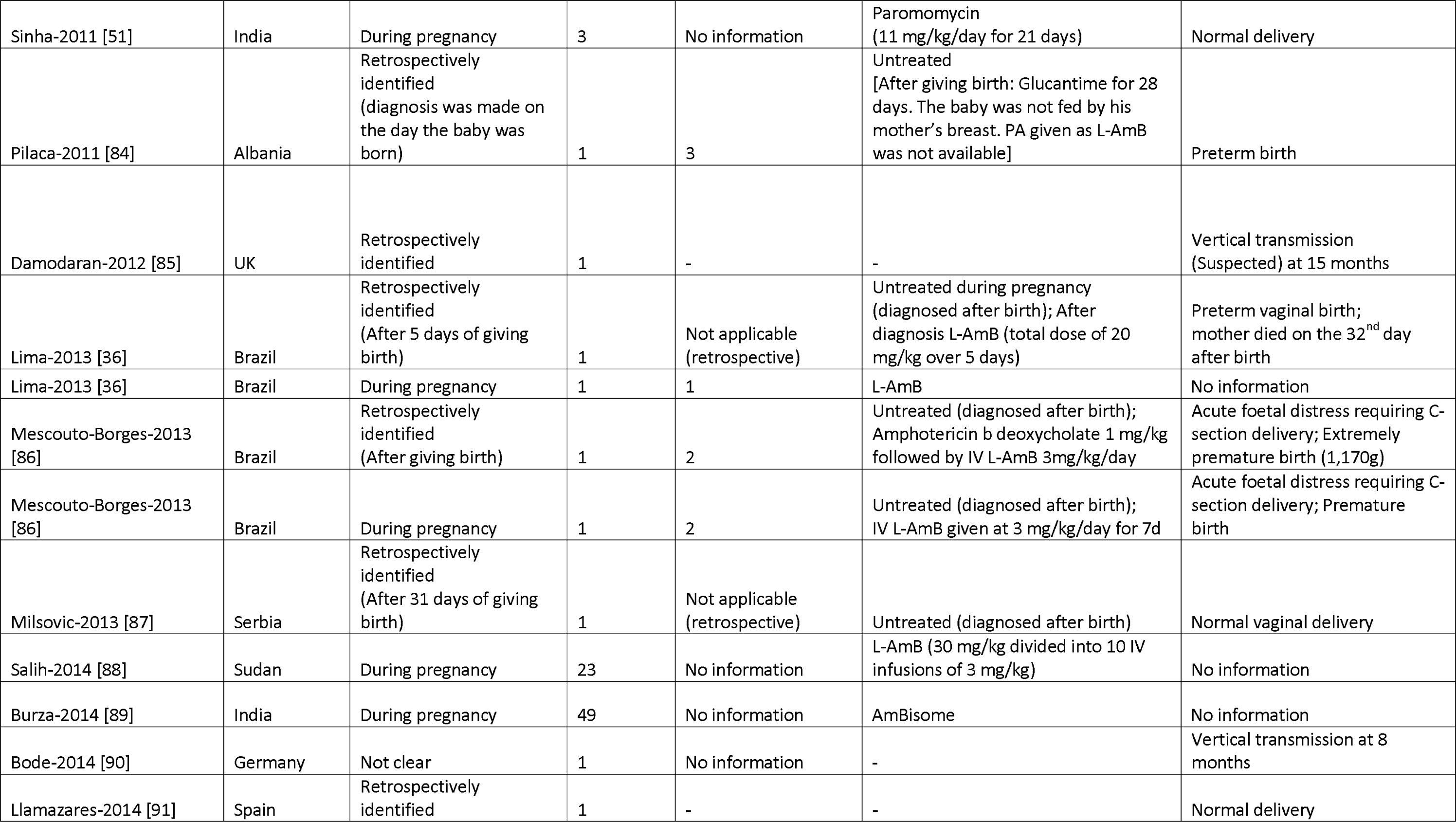
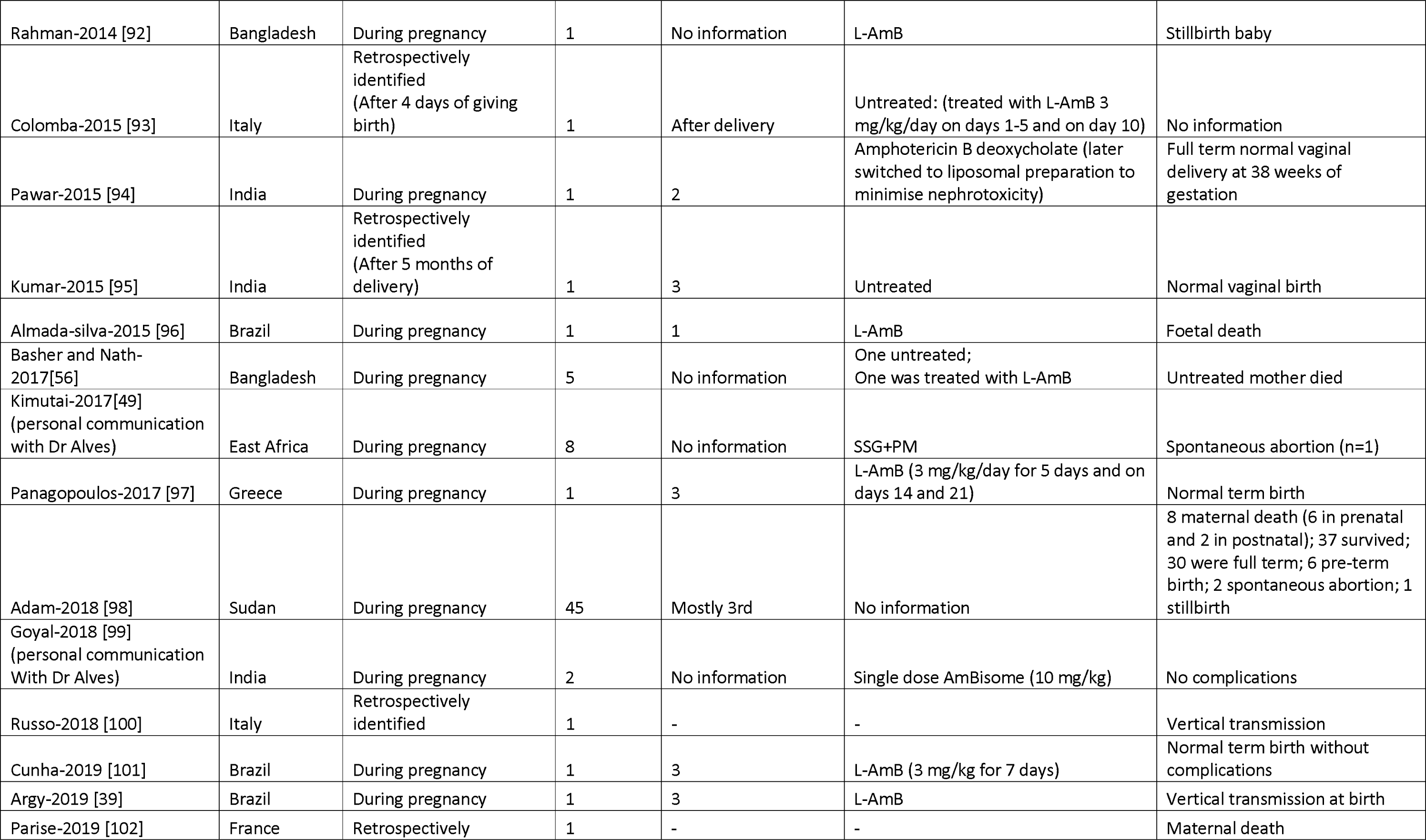
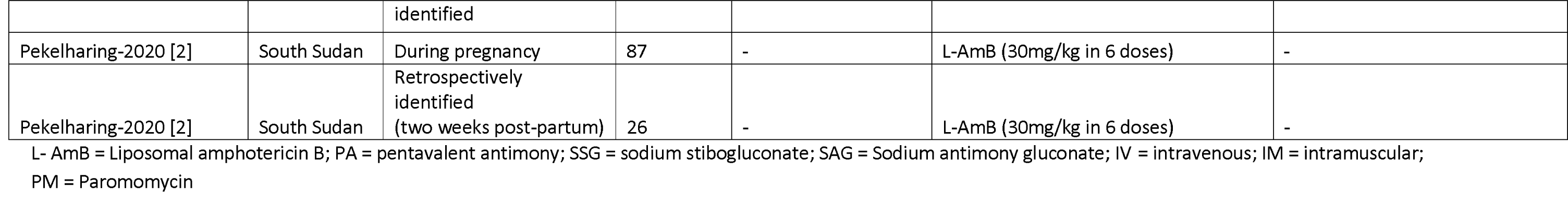
Description of reported 447 cases of VL in pregnancy

### Spatial distribution

A total of 21 (30.0%) records were from Europe, 21 (30.0%) from Southern Asia, 13 (18.6%) from South America, 8 (11.4%) from Northern America, 5 (7.1%) from Eastern Africa, and 1 (1.4%) record each were from Eastern and Western Asia. There were 16 records from India (22.8%), 13 (18.6%) from Brazil, 8 (11.4%) from Sudan, and further breakdown by country is presented in Fig 2 (left panel). There were 62 (88.6%) records in English language, 7 (10.0%) in Portuguese, and 1 (1.4%) in French. The 70 records included in this review described 447 cases of VL in pregnant women, of whom 159 (35.6%) were from Sudan, 113 (25.3%) from South Sudan, 80 (17.9%) from India, 23 (5.1%) from Bangladesh, 20 (4.5%) from Brazil, 12 (2.7%) from Italy, 10 (2.2%) from Uganda, and the rest of the breakdown is presented in Fig 2 (right panel).

**Fig 2:**
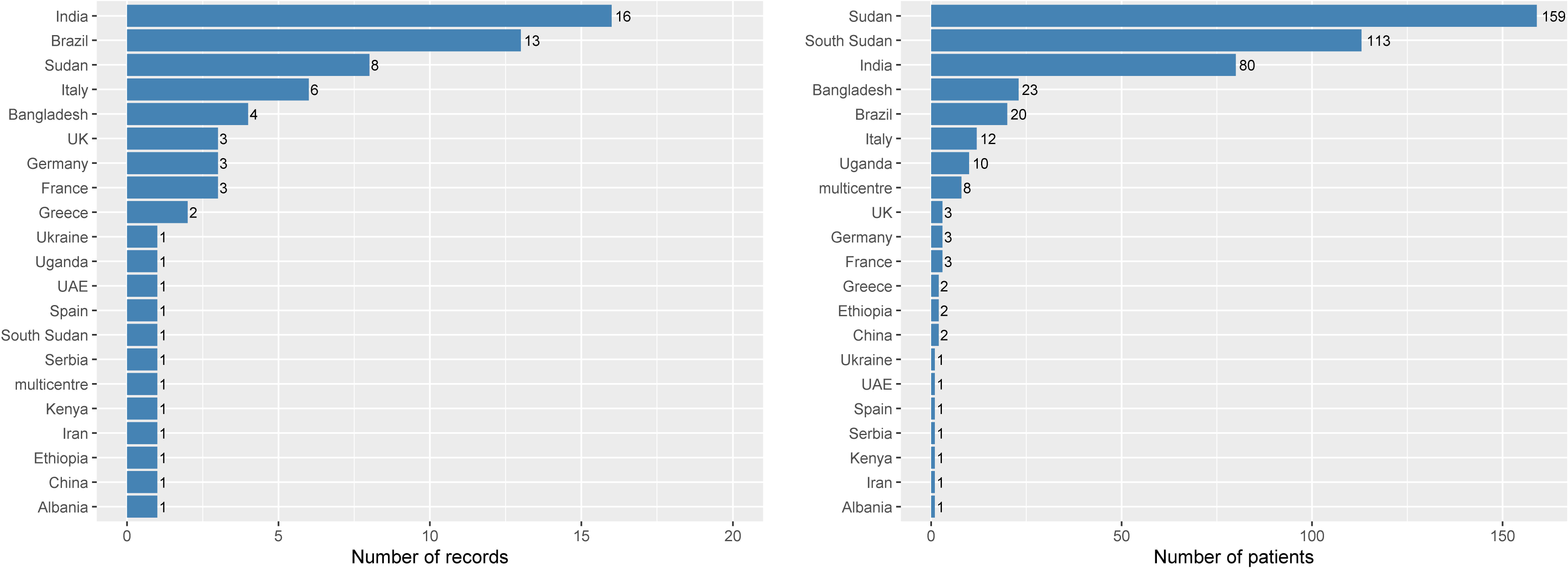
Number of records and patients by country of origin

### Treatment regimens

Of the 447 pregnant mothers identified, the disease was detected during pregnancy in 394 (88.1%), retrospectively confirmed after giving birth in 52 (11.6%), and the time of identification was not clear in one (0.2%). Ten (2.2%) were suspected of having carried the infection during their pregnancy, of whom 6 were cases of sub-clinical persistence of the parasites without the mother ever suffering from the disease previously. One case of oligosymptomatic mother was identified in Brazil [36] (Table 2). Of the 394 mothers whose infection was identified during pregnancy, 344 (89.1%) received a treatment, 3 (0.8%) were untreated, and the treatment status was not clear in the remaining 47 (12.2%) (Table 2). Description of characteristics and outcomes among 344 mothers who were treated and 3 untreated mothers are presented next.

Liposomal Amphotericin B (n=202) There were 5 (2.5%) mothers treated in the first trimester, 8 (4.0%) in second trimester, 9 (4.5%) in third trimester, and the time in pregnancy was not clear in 180 (89.1%). Survival status was not reported or was unclear in 26 (12.9%) mothers and from the remaining 176 mothers, a total of four (2.3%) maternal deaths were reported. There were a total of 5st (2.9%) miscarriages (trimester not clear), 1 (0.6%) foetal death (from a mother in 1^st^ trimester), 1 (0.6%) stillbirth (trimester not clear), and 1 (0.6%) premature birth (trimester not clear). Three cases of vertical transmission were identified [37–39]: one was detected immediately after vaginal birth (the baby was treated with L-AmB and recovered successfully), another at 11 months after birth (treatment information not available), and for the third case, vertical transmission was suspected at 8 months after birth (treated with sodium stibogluconate 20 mg/kg IV for 20 days and discharged) (Table 3).

**Table 3:**
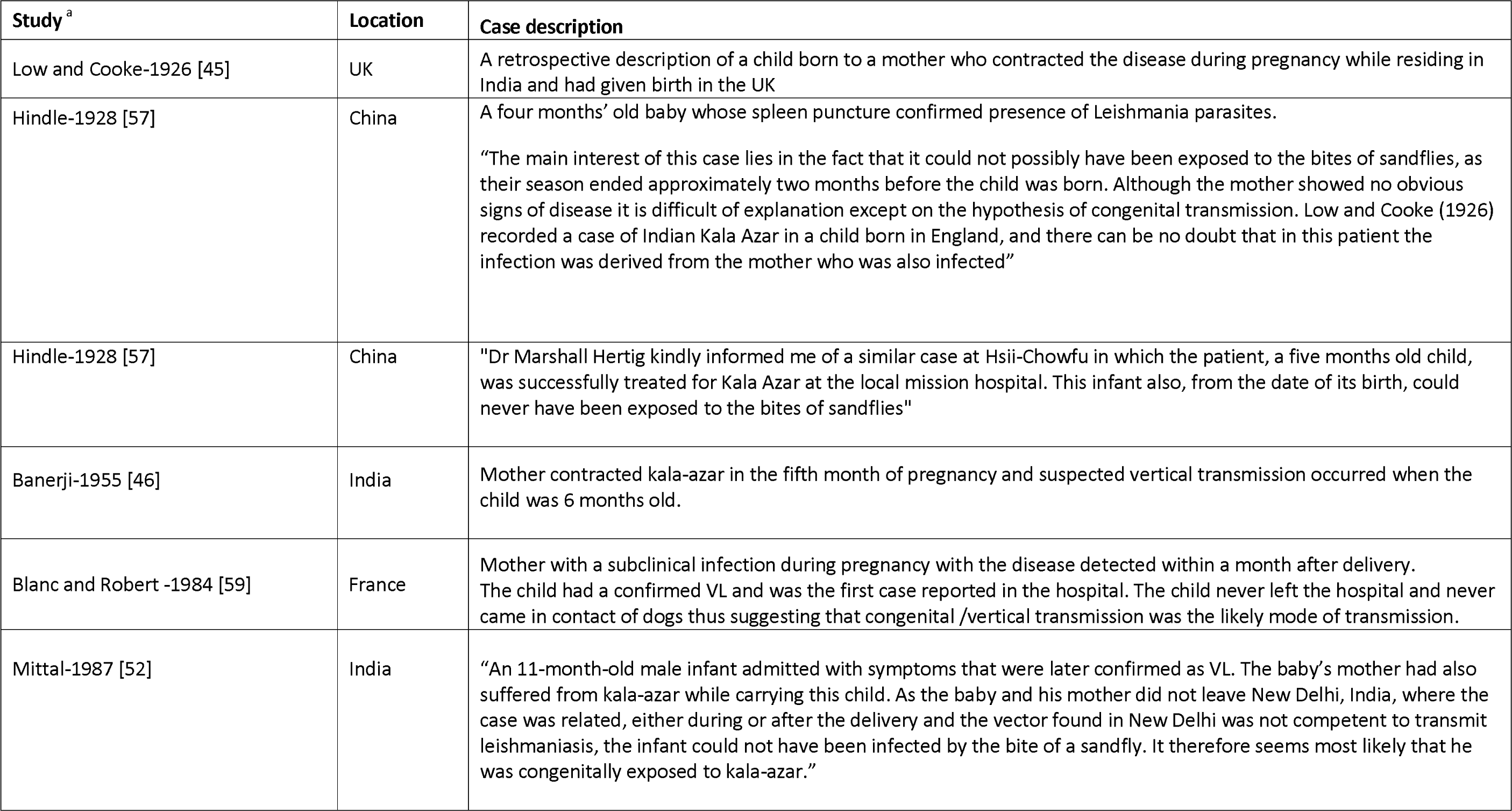
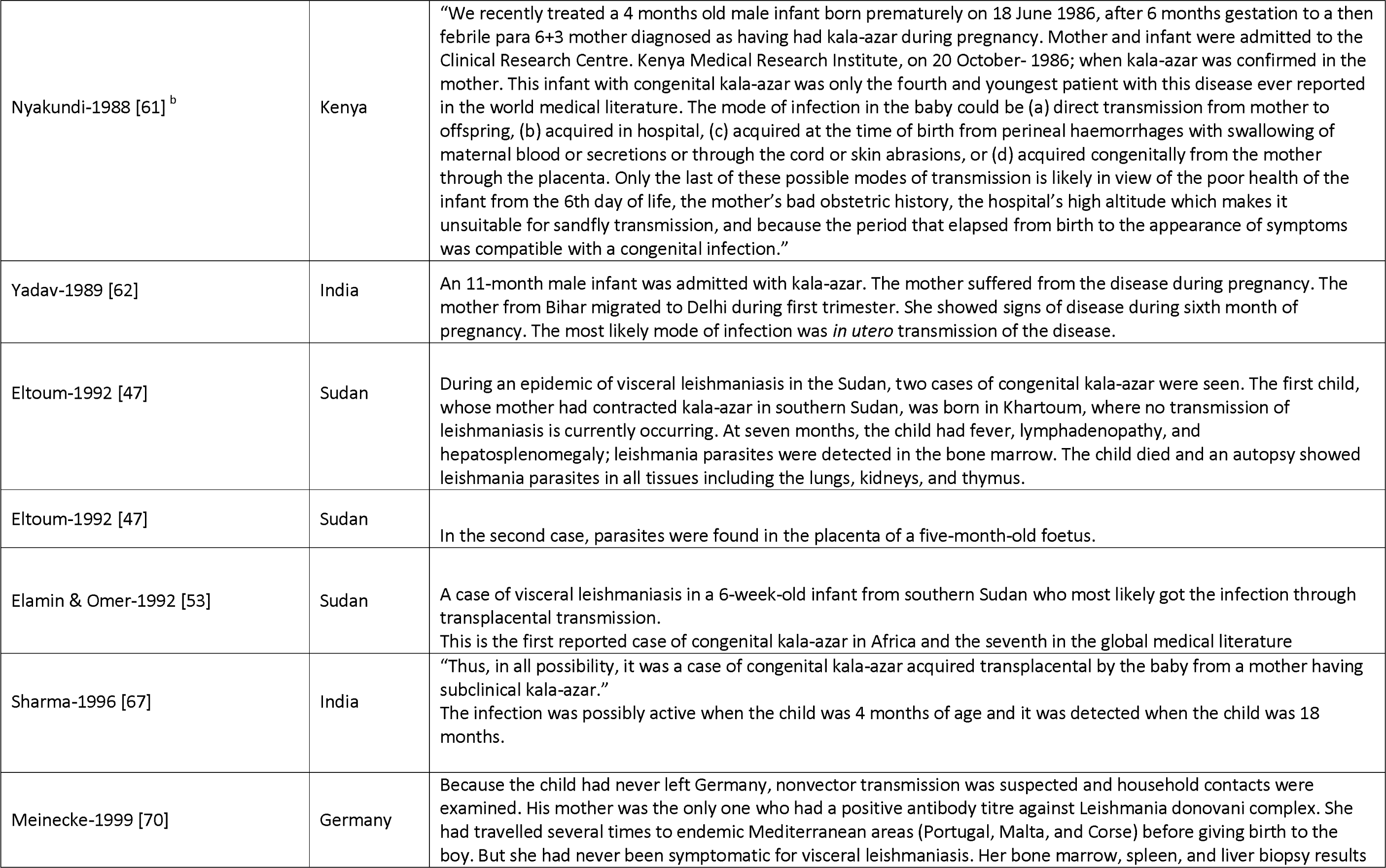
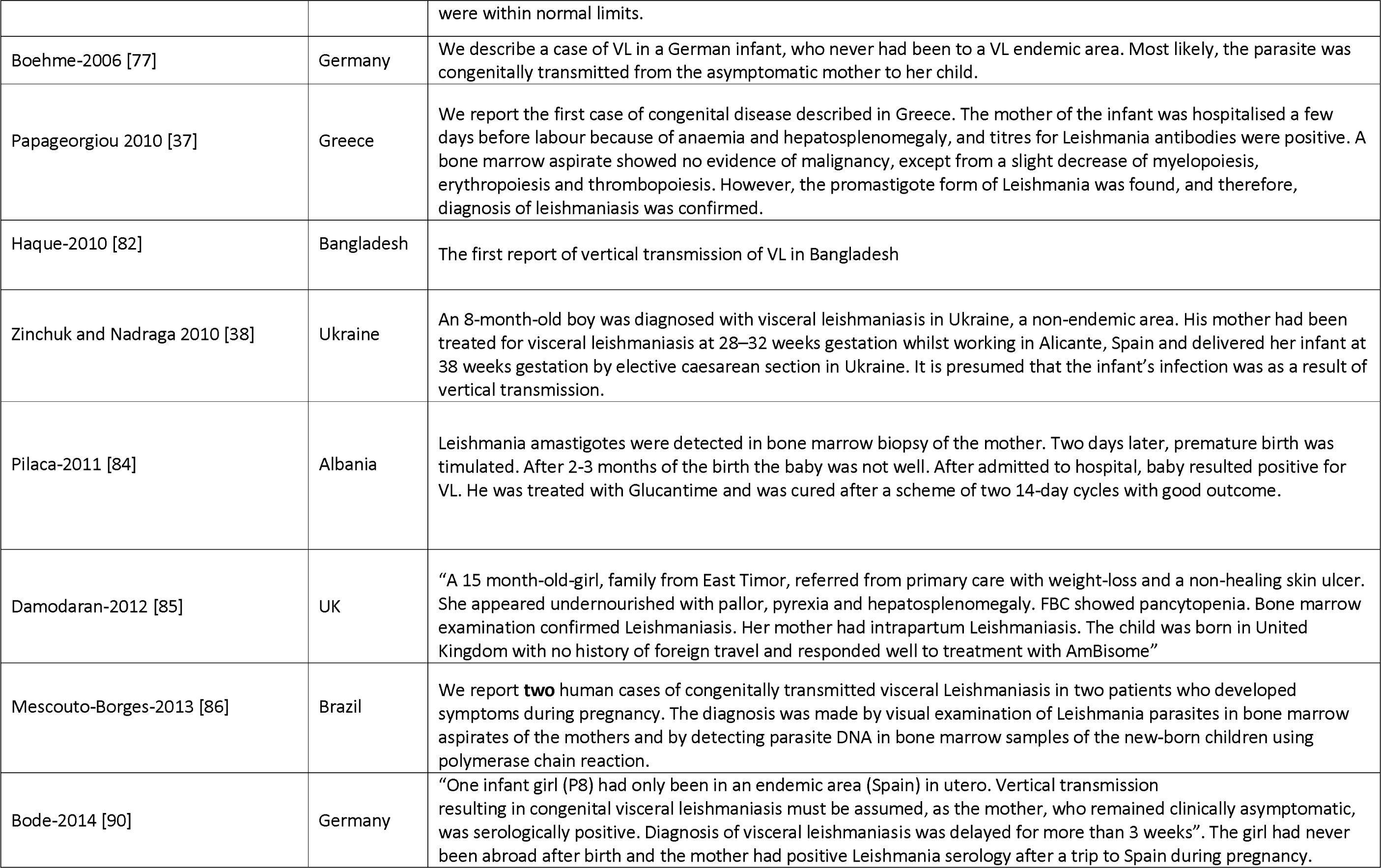
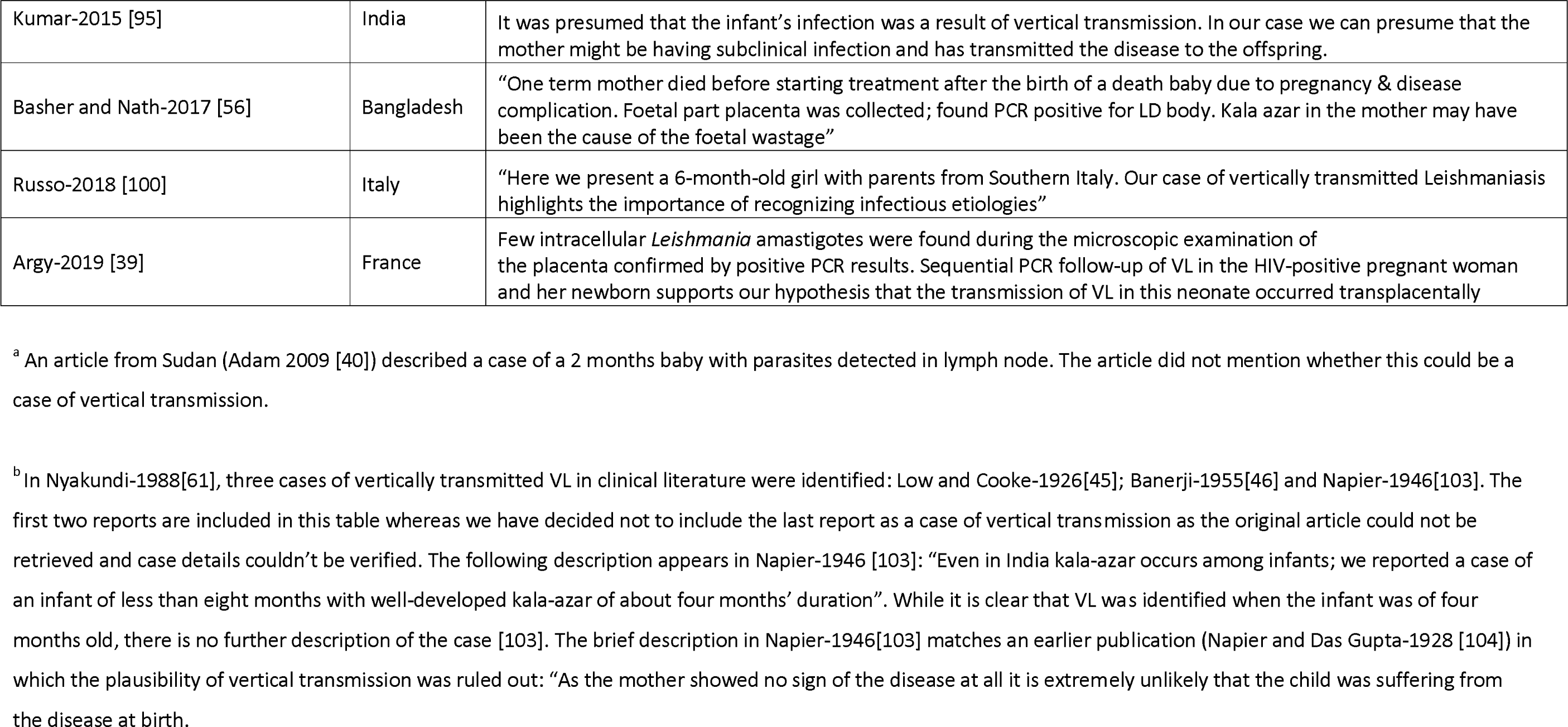
Details of 26 reported cases of confirmed or suspected congenital VL

### Pentavalent antimony (n=92)

There were 20 (21.7%) mothers in the first trimester, 45 (48.9%) in the second, 22 (23.9%) in the third, and the time in pregnancy was not clear in 5 (5.4%). Survival status was available on 87 (94.6%) mothers of whom 4 (4.6%, 4/87) died due to hepatic encephalopathy [40]. There were 24 (27.6%) abortions (or spontaneous abortions) [13, 41], 2 (2.3%) miscarriages, 2 (2.3%) pre-term births [40, 42], and 1 (1.1%) mother required splenectomy after delivery due to poor recovery [43]. One of the babies died due to myelomeningocele 3 hours after birth [40], another died one day after being born [44], another died due to VL at 2 months [40], and one was born with Down’s syndrome to a 47 years old mother [40]. There were 3 cases of vertical transmission identified [45–47], detected at 6, 7, and 12 months after birth. All three of them were treated with PA; 1 baby died (who was born with signs of intra- uterine growth retardation and was diagnosed with vertical VL at 7 months) and the other two survived (Table 3).

### Amphotericin B deoxycholate (n=20)

Of the 20 mothers treated with amphotericin B deoxycholate, 3 (15.0%) were in their first trimester, 6 (30.0%) in the second, 3 (15.0%) in the third, and the trimester was not clear in 8 (40.0%). There was one (5.0%) maternal death after 7 days of treatment due to haemorrhagic complications occurring after delivery (the mother was in 28.7 ± 7.8 weeks of pregnancy–exact time not available) [48]. The remaining 19 mothers were discharged alive.

The delivery of babies was described as normal for 18 mothers, haemorrhagic complication occurred in a mother after delivery (as described earlier) [48], and the information was not reported on 1. There was no evidence of vertical transmission of the disease in 12 (60.0%) babies in whom the information was reported. Nineteen of the children were alive and the survival status for 1 was missing.

### Pentavalent antimony plus paromomycin (aminosidine) (n=11)

Eleven pregnant mothers were treated with sodium stibogluconate plus aminosidine (paromomycin) [14, 49]. Information regarding trimester, maternal survival status, or vertical transmission were not available. One spontaneous abortion was reported [49].

### Liposomal amphotericin B plus pentavalent antimony (n=4)

Four mothers were treated with the combination regimen [50], of whom two in the second trimester and two in their third. L-AmB was administered at 3–7 mg/kg daily on days 1, 6, 11 and 16 (or on days 1, 2, 3, 4, 10 and 15), followed by 20 mg/kg sodium stibogluconate intramuscularly once daily for 30 days. All four mothers were discharged alive. At discharge, one mother delivered a healthy baby and the remaining three were still pregnant – no follow-up data was available.

### Paromomycin (aminosidine) (n=3)

Three pregnant mothers were treated with paromomycin administered by deep gluteal intramuscular injection once daily for 21 consecutive days [51]. The delivery was described as normal for all three with normal healthy babies at birth and all three mothers were alive.

### Unclear drug name (n=12)

Two publications described one case each without reporting the name of the drug administered [52, 53]. In an article, the number of mothers (n=10) allocated to each drug arm (pentavalent antimony or amphotericin B deoxycholate) was not clear [54]. Two (16.7%) of the mothers were in their third trimester and the status was unknown for the remaining 10 (83.3%). There were two (16.7%) maternal deaths [54] and two cases of vertical transmission [52, 53]. The first one was identified at 8 months after birth and another at 6 weeks after birth (the baby died after 3 days). Both babies were administered treatment upon detection of VL.

### Untreated (n=3)

Three cases of VL identified during pregnancy were untreated [12,55,56]. Treatment was deferred until after delivery due to safety concerns in one study [12]; there were signs of intra-uterine growth retardation requiring emergency C-section, and both mother and the child were alive. The second mother was not treated due to lack of adequate hospital resources [55]; the baby born to the mother died after 2 months due to malnutrition with no evidence of vertical transmission. In the third case, VL was diagnosed but the mother died after giving birth and before treatment could be administered [56]; the baby also died and foetal part placenta examination revealed presence of Leishman Donovan bodies by PCR indicating vertical transmission.

### Confirmed/probable/suspected vertical transmission

We identified a total of 26 cases of confirmed, probable or suspected cases of vertical transmission (Table 3). The median time to detect vertically transmitted VL was 6 months (range: 0–18 months). Eleven children were born to mothers in whom the disease status was confirmed during their pregnancy (3 were treated with L-AmB, 3 were treated with PA, the drug name was not clear in 2, 1 was untreated and the treatment status was not clear in remaining 2). Histopathological examination of the placenta confirmed the vertical transmission of the disease in two cases [47, 56] and this was not reported for the remaining cases. Treatment status was described in 18 children, of whom 11 received pentavalent antimony, 6 received L-AmB and 1 received amphotericin b deoxycholate. Two of the children died (one received pentavalent antimony and the drug name was not clear in the other).

### Risk of bias assessment

Of the 69 case reports/case series, 48 (69.6%) were considered to be at a high risk of bias in patient selection, 9 (13.0%) were at high risk of exposure (confirmed VL status) ascertainment bias, 12 (17.4%) were at high risk of outcome ascertainment bias, 13 (18.8%) at high risk of incomplete reporting bias, and 14 (20.3%) studies were at a high risk of bias due to inadequate follow-up (See S1 Table). One retrospective cohort study with a comparative group of non-pregnant patient group was considered of high quality.

## Discussion

The occurrence and effects of VL during pregnancy is under-researched and poorly understood as evidenced by having identified only 70 publications describing a total of 447 cases of VL in pregnancy in the past 90 years.

The small case volume reported in the literature could have several explanations. In the first place, there is an apparent imbalance in caseloads with predominance of the disease among males; ascribed to biological or behavioural causes [3,17,63,105–107]. Pentavalent antimony is contraindicated in pregnancy and was the first line therapy before the development of Liposomal amphotericin B (L-AmB) – this might have traditionally dissuaded physicians from treating VL during pregnancy and leaning towards postponing the treatment until after delivery unless treatment is absolutely warranted [12]. However, this situation might have changed recently as liposomal amphotericin B has no contraindication during pregnancy and is the treatment of choice. It has also been postulated that early pregnancies are missed due to spontaneous abortion caused by VL [63]. Women with childbearing potential or those who are already pregnant are systematically excluded from VL clinical studies and only a third of the patients enrolled in clinical trials are females [17]. For example, of the 158 studies indexed (to date) in the IDDO systematic library of VL clinical trials, 52 studies presented details from screening logs of patients (33,455 patients were screened; 17,572 patients were excluded including 32 pregnant women, and 15,883 included) (See S1 Data) [32]. Assuming all those screened for eligibility indeed had the disease, this would give an estimated 0.096% (likely an underestimate) of the total cases of VL to be pregnant women. If there are 100,000 cases per year, this translates to a minimum of 96 cases of the disease in pregnancy per year. It is clear that the likely size of the problem is much bigger than what can be estimated from available reports. For example, during Jan 2016–Jul 2019 in Lankien, Jonglei state, South Sudan, out of 4,448 cases of VL diagnosed, 39% occurred in women of childbearing age, and 13% of the women (2.5% of all cases) were pregnant [2]. It is also likely that the clustering in space and time of reported cases (more than half of all cases in this review were from studies in Sudan or South Sudan after 2005) is more a result of local interest into the subject matter than a true reflection of disease burden.

There was also geographical disparity in the treatment regimens used, reflecting heterogeneity in treatment practices. Only half of the patients received amphotericin B regimens in studies conducted in Africa compared to more than two-thirds of the patients from Asia. There was a total of 11 maternal deaths; four (4.6%, 4/87) occurred in those treated with pentavalent antimony-based regimens, 4 (4/176; 2.3%) among those treated with L-AmB, 1 (1/20, 5.0%) with amphotericin b deoxycholate, and the drug name used for the treatment was not clear in 2 (16.7%,2/12) cases. Spontaneous abortion following PA regimen was observed in just over a quarter of the mothers (24/88, 27.3%) while there were a total of 5 (2.9%) miscarriages and 1 (0.6%) foetal death following L-AmB regimen. Taken together, these results support the use of liposomal amphotericin B for the treatment of VL during pregnancy.

Our review identified 26 cases of vertically transmitted VL with a median time of detection of 6 months (range: 0–18 months). This suggests that children born to mothers with VL during pregnancy require a longer post-treatment follow-up than the standard 6-months follow-up duration among non-pregnant patients to monitor the well-being of the maternal-foetal pair. The underlying mechanism of the onset of clinical leishmaniasis among neonates and infants born to a successfully-treated mother during pregnancy (2.4% overall) is currently not clear; it has been ascribed to imbalances in immune-mechanism modulated by T cell responses (Th1/Th2) [10] or by parasites entering a state of dormancy in the lymph nodes [72].

Our review has identified limitations in reports of VL in pregnancy. Complete information was often not available on treatment administered and on efficacy and safety outcomes for the mother and baby. For 12% of the mothers, it could not be ascertained whether they had received any treatment or not. Majority of the studies were considered to be a high risk of bias for patient selection while some retrospective studies were at high risk of bias for ascertainment of exposure domain as VL diagnosis was purely based on clinical signs and symptoms or suspicion. This suggests that existing practices for management of VL in pregnancy is guided by limited evidence generated from case reports and small case series. High quality studies (such as Pekelharing-2020 [2]) is warranted for generation of a robust evidence regarding safety and efficacy of antileishmanial agents during pregnancy. There was also a lack of standardised reporting as information was missing on several critical parameters such as trimester status, time on detection of VL, and therapeutic outcomes of the mother and the child. Taken together, these findings highlight the need to improve and harmonise the reporting of VL in pregnant women. We have outlined a minimum checklist of items that might be useful for reporting purposes (Box 1).

As conducting randomised controlled trials during pregnancy poses ethical challenges, it is important to maximise currently available information from observational studies and case reports to gauge the potential safety of the therapies in pregnant women. Data from mothers who become pregnant after completion of therapy but within the follow-up period enrolled in trials might provide further resource, especially on the reproductive consequences of the treatment (Table 4). The recently proposed safe ethical framework for the recruitment of women susceptible to and becoming pregnant is an important development towards filling the existing knowledge gap [18]. Like for many NTDs, there is currently an absence of a comprehensive pregnancy-specific registry for exposures to antileishmanials, with the exception of the one dedicated for miltefosine [108]. Therefore, creating an open registry where all these cases are indexed and continually updated would help in better characterisation of the safety aspects of the drugs. Finally, the Infectious Diseases Data Observatory (IDDO) data platform, that is currently standardising individual participant data from several VL clinical studies, offers a unique resource to explore host, parasite, and drug dynamics affecting the safety and efficacy in pregnant populations [109].

**Table 4:** Description of patients enrolled in clinical trials who became pregnant after completion of treatment

| Study | Number of patients | Treatment received at enrolment | Pregnancy and outcome description |
| --- | --- | --- | --- |
| Bhattacharya-2007 [110] | 2 | Miltefosine | "Despite extensive counselling for contraception, 2 cases of pregnancy were reported, with the conception date close to the exposure period. One patient became pregnant 2 weeks after the end of treatment, and the other became pregnant at 3 months after the end of the treatment period. Two healthy babies were delivered at gestational weeks of 39 and 40, without any birth anomaly" |
| Sinha-2011 [51] | 1 | Paromomycin | One female patient became pregnant more than 1 month after completing treatment. The offspring was born alive and determined to be normal/healthy just after birth. |
| Mondal-2014 [111] | 4 | Liposomal Amphotericin B (Single dose) | Four female participants became pregnant within months after treatment and in one the pregnancy was completed with delivery of a term normal birth after 6 months of follow-up. The other three pregnant women were clinically healthy during the last follow-up visit. |
| Jamil-2015 [112] | 1 | Paromomycin | Pregnancy was reported in one female during the follow-up period. The offspring was born healthy, and a hearing test conducted on the infant at 1.5 months of age confirmed reaction to sound. An otoscopy and oto-acoustic emission test to determine function of the middle and inner ear was conducted at 3 months of age and confirmed normal hearing function. |
| Pandey-2016[113] | 15 | Miltefosine | All the female patients were suggested not to become pregnant within 6 months of treatment. However, 15 patients became pregnant within 6 months of follow-up (after 2 months of treatment completion). All these patients became pregnant 2 months after end of treatment. All of them were followed up till 1 year and all had full-term normal pregnancy with no congenital anomalies. |

## Conclusions

In conclusion, this review brings together scattered observations on VL in pregnant women and the cases of vertically transmitted VL reported in the clinical literature. Available reports clearly underestimate the scale of the problem. Existing therapeutic guidelines regarding the usage of drugs in pregnancy is guided by limited evidence generated from case reports and small case series. Our review suggests that liposomal amphotericin B should be the preferred treatment for VL during pregnancy.

#### List of boxes

**Box 1:**
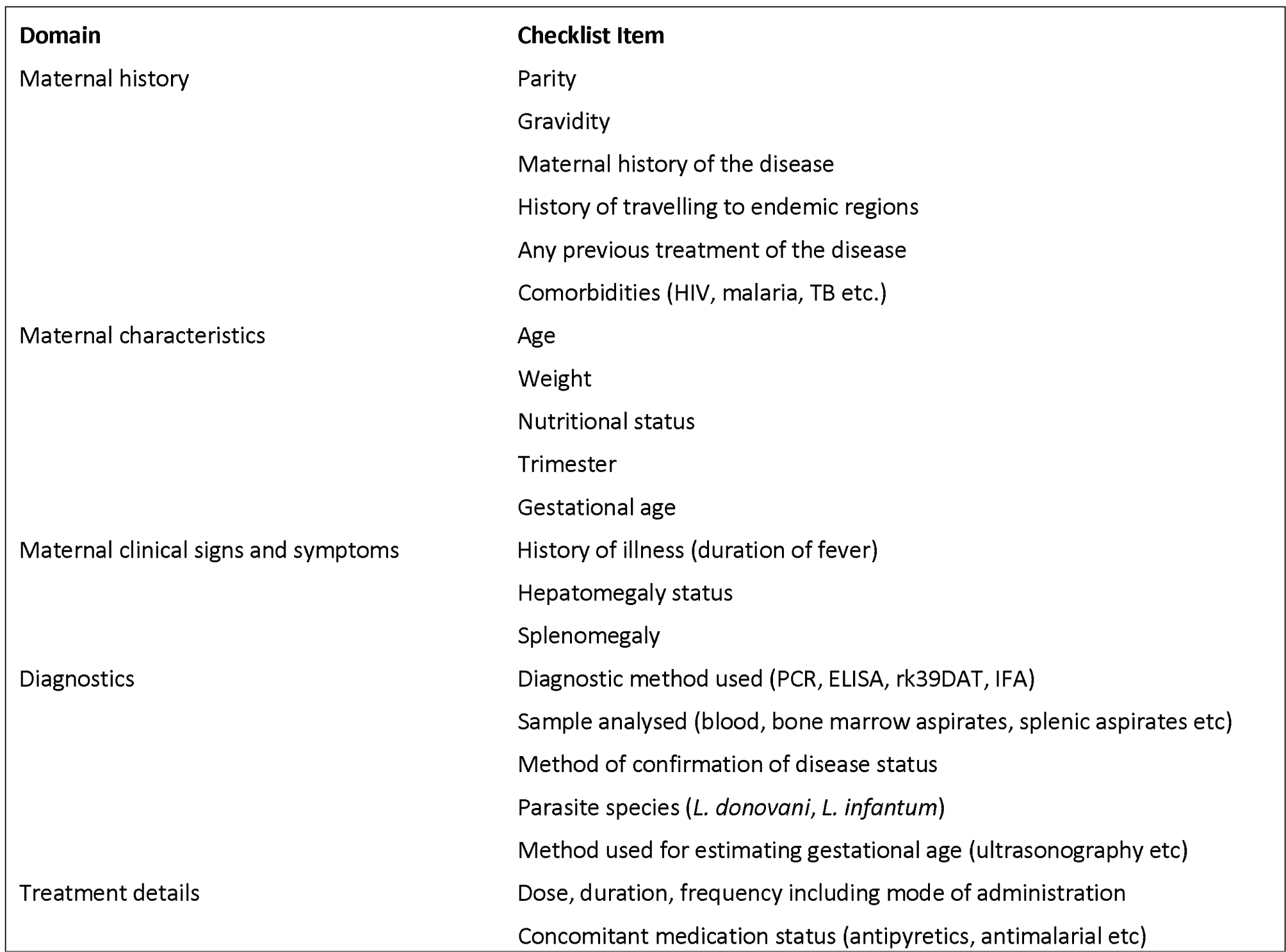
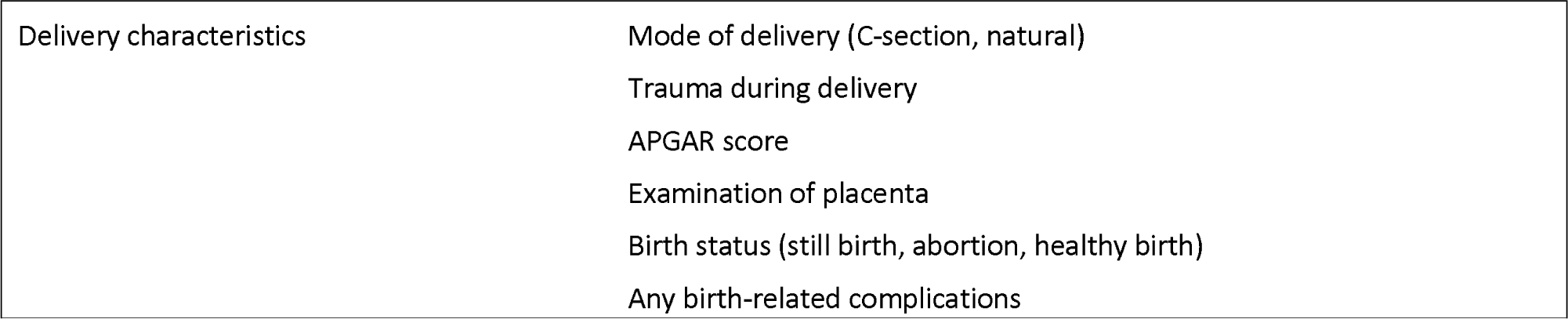
Proposed minimum variable recording and reporting for studies or case reports for VL in pregnancy

**Box 1: Proposed minimum variable recording and reporting for studies or case reports for VL in pregnancy** **Adapted from Saito-2017 [114]**
| Domain | Checklist Item |
| --- | --- |
| Maternal history | Parity<br>Gravidity<br>Maternal history of the disease<br>History of travelling to endemic regions<br>Any previous treatment of the disease<br>Comorbidities (HIV, malaria, TB etc.) |
| Maternal characteristics | Age<br>Weight<br>Nutritional status<br>Trimester<br>Gestational age |
| Maternal clinical signs and symptoms | History of illness (duration of fever)<br>Hepatomegaly status<br>Splenomegaly |
| Diagnostics | Diagnostic method used (PCR, ELISA, rk39DAT, IFA)<br>Sample analysed (blood, bone marrow aspirates, splenic aspirates etc)<br>Method of confirmation of disease status<br>Parasite species (*L. donovani*, *L. infantum*)<br>Method used for estimating gestational age (ultrasonography etc) |
| Treatment details | Dose, duration, frequency including mode of administration<br>Concomitant medication status (antipyretics, antimalarial etc) |
| Delivery characteristics | Mode of delivery (C-section, natural) |
|  | Trauma during delivery |
|  | APGAR score |
|  | Examination of placenta |
|  | Birth status (still birth, abortion, healthy birth) |
|  | Any birth-related complications |

## Data Availability

The database(s) supporting the conclusions of this article are available within the tables and figures presented within the manuscript along with the supplemental files (S1 Data, S2 Data).

## Declarations

### Authors’ contributions

Conceptualization : PD, PJG, PLO

Data Curation : PD, SSP

Formal Analysis : PD, SSP, PJG, PLO

Funding Acquisition : PJG

I vestignation : PD, SSP, PJG, PLO

Methodology : PD, SSP, EH, BJM

Project Administration : PD, PLO

Resources : PJG, PLO

Software : PD

Supervision : PJG, PLO

Validation : PD, SSP, PJG, PLO

Visualization : PD

Writing – Original Draft Preparation : PD, PLO

Writing – Review & Editing : PD, SSP, BJM, EH, KR, FA, PJG, PLO

### List of supplemental files

S1 Text: PRISMA checklist

S2 Text: Search details

S1 Data: Screening list

S2 Data: Study data

S1 Table: Risk of bias assessment

### Ethics approval and consent to participate

Not applicable

### Consent for publication

Not applicable

## Acknowledgements

We would like to thank for the Prof. Bernhard Lämmle and his team for helpful responses on queries related to their manuscript.

## References

1. WHO. Leishmaniasis: Key facts [Internet]. WHO. 2020 [cited 2020 Jul 5]. Available from: https://www.who.int/news-room/fact-sheets/detail/leishmaniasis

2. Pekelharing JE, Gatluak F, Harrison T, Maldonado F, Siddiqui R, Ritmeijer K. Outcomes of visceral leishmaniasis in pregnancy : A retrospective cohort study from South Sudan. PLoS Negl. Trop. Dis. 2020;14:e0007992.

3. World Health Organization. Control of the leishmaniases. World Heal. Organ. Tech. Rep. Ser. 949. 2010;22–6.

4. Alvar J, Croft S, Olliaro P. Chemotherapy in the Treatment and Control of Leishmaniasis. Adv. Parasitol. 2006;61:223–74.

5. Sundar S, Olliaro PL. Miltefosine in the treatment of leishmaniasis: Clinical evidence for informed clinical risk management. Ther. Clin. Risk Manag. 2007;3:733–40.

6. Boelaert M, Sundar S. Leishmaniasis. In: Farrar J, Hotez PJ, Junghanss T, Kang G, Lalloo D, White NJ, editors. Manson’s Trop. Dis. 23rd ed. Elsevier Health Sciences; 2013. p. 631–51.

7. Nuwayri-salti N, Khansa HF. Direct non-insect-vector transmission of leishmania parasites in mice. Int. J. Parasitol. 1985;15:497–500.

8. Utili R, Rambaldi A, Tripodi MF, Andreana A. Visceral leishmaniasis during pregnancy treated with meglumine antimoniate. Infection. 1995;23:182–3.

9. Pagliano P, Carannante N, Rossi M, Gramiccia M, Gradoni L, Faella FS, et al. Visceral leishmaniasis in pregnancy: A case series and a systematic review of the literature. J. Antimicrob. Chemother. 2005;55:229–33.

10. Berger BA, Bartlett AH, Saravia NG, Galindo Sevilla N. Pathophysiology of Leishmania Infection during Pregnancy. Trends Parasitol. 2017;33:935–46.

11. Pagliano P, Ascione T, Di Flumeri G, Boccia G, De Caro F. Visceral leishmaniasis in immunocompromised: Diagnostic and therapeutic approach and evaluation of the recently released IDSA guidelines. Infez. Med. 2016;24:265–71.

12. Kumar A, Mittal M, Prasad S. Treatment of leishmaniasis in pregnancy. Int. J. Gynecol. Obstet. 2001;72:189–90.

13. Miah M, Ayaz F, Maniruzzaman M, Ahasan M, Bari S, Mawla S, et al. Kala azar in pregnancy. Mymensingh Med. J. 2010;Oct; 19:529–32.

14. Seaman J, Pryce D, Sondorp HE, Moody A, Bryceson ADM, Davidson RN. Epidemic Visceral Leishmaniasis in Sudan: A Randomized Trial of Aminosidine plus Sodium Stibogluconate versus Sodium Stibogluconate Alone. J. Infect. Dis. 1993;168:715–20.

15. Kumar D, Ramesh V, Verma S, Ramam M, Salotra P. Post-kala-azar dermal leishmaniasis (PKDL) developing after treatment of visceral leishmaniasis with amphotericin B and miltefosine. Ann. Trop. Med. Parasitol. 2009;103:727–30.

16. Verma P, Grover C, Sharma S. Post-kala-azar dermal leishmaniasis in pregnancy: Hitherto unaccounted. Int. J. Dermatol. 2014;53:1501–4.

17. Dahal P, Singh-Phulgenda S, Olliaro PL, Guerin PJ. Gender disparity in patients enrolled in clinical trials of visceral leishmaniasis: a systematic review and meta-analysis. PLoS Negl. Trop. Dis. 2021;[In Press].

18. Couderc-Pétry M, Eléfant E, Wasunna M, Mwinga A, Kshirsagar NA, Strub-Wourgaft N. Inclusion of women susceptible to and becoming pregnant in preregistration clinical trials in low-and middle-income countries: A proposal for neglected tropical diseases. PLoS Negl. Trop. Dis. 2020;14:1–15.

19. Banjara MR, Hirve S, Siddiqui NA, Kumar N, Kansal S, Huda MM, et al. Visceral leishmaniasis clinical management in endemic districts of India, Nepal, and Bangladesh. J. Trop. Med. 2012;2012.

20. Treatment of Leishmaniasis With Impavido® (Miltefosine): Pregnancy Registry [Internet]. [cited 2021 Feb 9]. Available from: https://clinicaltrials.gov/ct2/show/NCT02427308

21. IMPAVIDO (miltefosine) Pregnancy Registry [Internet]. [cited 2021 Feb 9]. Available from: https://www.impavido.com/about-registry

22. Figueiró-Filho EA, Duarte G, El-Beitune P, Quintana SM, Maia TL. Visceral leishmaniasis (kala-azar) and pregnancy. Infect. Dis. Obstet. Gynecol. 2004;12:31–40.

23. Silva JSF e., Galvao TF, Pereira MG, Silva MT. Treatment of American tegumentary leishmaniasis in special populations: A summary of evidence. Rev. Soc. Bras. Med. Trop. 2013;46:669–77.

24. Kip AE, Schellens JHM, Beijnen JH, Dorlo TPC. Clinical Pharmacokinetics of Systemically Administered Antileishmanial Drugs. Clin. Pharmacokinet. 2018;57:151–76.

25. The electronic Medicines Compendium. Pentostam Injection [Internet]. 2019 [cited 2019 Aug 30]. Available from: https://www.medicines.org.uk/emc/product/5466/smpc

26. World Health Organization. WHO Technical Report Series 949: Control of the leishmaniases. 2010.

27. The electronic Medicines Compendium. AmBisome [Internet]. 2019 [cited 2019 Aug 30]. Available from: https://www.medicines.org.uk/emc/product/1022#PREGNANCY

28. WHO. Guidelines for diagnosis, treatment and prevention of visceral leishmaniasis in South Sudan [Internet]. WHO. [cited 2019 Oct 10]. Available from: https://www.who.int/leishmaniasis/burden/Guidelines_for_diagnosis_treatment_and_prevention_of_VL_in_South_Sudan.pdf

29. FDA. IMPAVIDO (miltefosine) prescribing information [Internet]. www.accessdata.fda.gov. [cited 2019 Oct 10]. Available from: https://www.accessdata.fda.gov/drugsatfda_docs/label/2014/204684s000lbl.pdf

30. Ministry of Health R of K. Prevention, Diagnosis and Treatment of Visceral Leishmaniasis (Kala-Azar) in Kenya [Internet]. 2017 [cited 2019 Aug 30]. Available from: https://www.who.int/leishmaniasis/burden/Kala_Azar_Kenya_2017.pdf?ua=1

31. Moher D, Liberati A, Tetzlaff J, Altman DG. Reprint--preferred reporting items for systematic reviews and meta-analyses: the PRISMA statement. PLoS Med. 2009;6:e1000097.

32. Infectious Diseases Data Observatory. VL Surveyor [Internet]. www.iddo.org. 2020 [cited 2020 Sep 17]. Available from: https://www.iddo.org/vlSurveyor/#0

33. United Nations. Standard country or area codes for statistical use [Internet]. [cited 2018 Sep 13]. Available from: https://unstats.un.org/unsd/methodology/m49/overview/

34. R Core Team. R: A language and environment for statistical computing. R Found. Stat. Comput. Vienna, Austria. 2018.

35. Murad MH, Sultan S, Haffar S, Bazerbachi F. Methodological quality and synthesis of case series and case reports. Evid. Based. Med. 2018;23:60–3.

36. Lima TB. Liver biopsy for visceral leishmaniasis diagnosis in pregnancy: report of 2 cases. World J. Clin. Infect. Dis. 2013;3:20–4.

37. Papageorgiou T, Pana Z, Tragiannidis A, Tsotoulidou V, Pratsiou E, Tzouvelekis G, et al. The first case of congenital leishmaiasis in a female infant in Greece. J. Paediatr. Child Health. 2010;46:611–2.

38. Zinchuk A, Nadraga A. Congenital visceral leishmaniasis in Ukraine: case report. Ann. Trop. Paediatr. 2010;30:161–4.

39. Argy N, Lariven S, Rideau A, Lemoine A, Bourgeois Moine A, Allal L, et al. Congenital Leishmaniasis in a Newborn Infant Whose Mother was Coinfected With Leishmaniasis and HIV. J. Pediatr. Infect. Dis. Soc. 2019;07:7.

40. Adam GK, Abdulla MA, Ahmed AA, Adam I. Maternal and perinatal outcomes of visceral leishmaniasis (kala-azar) treated with sodium stibogluconate in eastern Sudan. Int. J. Gynecol. Obstet. 2009;107:208–10.

41. Mueller M, Balasegaram M, Koummuki Y, Ritmeijer K, Santana MR, Davidson R. A comparison of liposomal amphotericin B with sodium stibogluconate for the treatment of visceral leishmaniasis in pregnancy in Sudan. J. Antimicrob. Chemother. 2006;58:811–5.

42. Silveira BP, Araujo Sobrinho J, Leite LF, Sales M, Gouveia Mdo S, Mathias RL, et al. [Premature birth after the use of pentavalent antimonial: case report]. Rev. Soc. Bras. Med. Trop. 2003;36:523–5.

43. El-Saaran AM. Visceral Leishmaniasis in Dubai. Trans R Soc Trop Med Hyg. 1979;73:475.

44. Silveira BP, Sobrinho JA, Leite LF, Andrade Sales M das N, Araújo Gouveia M do S, Mathias RL, et al. Parto prematuro após uso de antimonial pentavalente: Relato de um caso. Rev. Soc. Bras. Med. Trop. 2003;36:523–5.

45. Low GC, Cooke WE. A congenital case of kala-azar. Lancet. 1926;208:1209–11.

46. Banerji D. Possible congenital infection of kalaazar. J. Indian Med. Assoc. 1955;24.

47. Eltoum IA, Zijlstra EE, Ali MS, Ghalib HW, Satti MMH, Eltoum B, et al. Congenital kala- azar and leishmaniasis in the placenta. Am. J. Trop. Med. Hyg. 1992;46:57–62.

48. Figueiro-Filho EA, El Beitune P, Queiroz GT, Somensi RS, Morais NO, Dorval ME, et al. Visceral leishmaniasis and pregnancy: analysis of cases reported in a central-western region of Brazil. Arch. Gynecol. Obstet. 2008;278:13–6.

49. Kimutai R, Musa AM, Njoroge S, Omollo R, Alves F, Hailu A, et al. Safety and Effectiveness of Sodium Stibogluconate and Paromomycin Combination for the Treatment of Visceral Leishmaniasis in Eastern Africa: Results from a Pharmacovigilance Programme. Clin. Drug Investig. 2017;37:259–72.

50. Mueller M, Balasegaram M, Koummuki Y, Ritmeijer K, Santana MR, Davidson R. A comparison of liposomal amphotericin B with sodium stibogluconate for the treatment of visceral leishmaniasis in pregnancy in Sudan. J. Antimicrob. Chemother. 2006;58:811–5.

51. Sinha PK, Jha TK, Thakur CP, Nath D, Mukherjee S, Aditya AK, et al. Phase 4 Pharmacovigilance Trial of Paromomycin Injection for the Treatment of Visceral Leishmaniasis in India. J. Trop. Med. 2011;2011:1–7.

52. Mittal V, Sehgal S, Yadav T, Singh VK. Congenital transmission of kala-azar. J Commun Dis. 1987;Jun:184–5.

53. Elamin A, Omer MIA. Visceral Leishmaniasis in a 6-Week-Old Infant: Possible Congenital Transmission. Trop. Doct. 1992;22:133–5.

54. Mueller Y, Mbulamberi DB, Odermatt P, Hoffmann A, Loutan L, Chappuis F. Risk factors for in-hospital mortality of visceral leishmaniasis patients in eastern Uganda. Trop. Med. Int. Heal. 2009;14:910–7.

55. Vieira ML, Jacobina RR, Soares NM. Leishmaniose visceral em adolescente gestante. Rev. ciênc. méd. biol. 2007;6:357–61.

56. Basher A, Nath PN. Transplacental transmission of visceral Leishmaniasis; Looking for the evidence-A case series. Trop. Med. Int. Heal. 2017;22 (Supp):154.

57. Hindle E. Further Observations on Chinese Kala Azar. Proc. R. Soc. B Biol. Sci. 1928;103:599–619.

58. Rees PH, Kager PA, Wellde BT, Hockmeyer WT. The response of Kenyan kala-azar to treatment with sodium stibogluconate. Am. J. Trop. Med. Hyg. 1984;33:357–61.

59. Blanc C, Robert A. [5th case of congenital kala-azar]. Press. Med. 1984;Jul 7:1751.

60. Badaró R, Rocha H, Carvalho EM, Queiroz AC, Jones TC. Leishmania Donovani: an Opportunistic Microbe Associated With Progressive Disease in Three Immunocompromised Patients. Lancet. 1986;327:647–9.

61. Nyakundi PM, Muigai R, Were JBO, Oster CN, Gachihi GS, Kirigi G. Congenital visceral leishmaniasis: Case report. Trans. R. Soc. Trop. Med. Hyg. 1988;82:564.

62. Yadav T, Gupta H, Satteya U, Kumar R, Mittal V. Congenital kala-azar. Ann Trop Med Parasitol. 1989;Oct:535–7.

63. Thakur C, Sinha G, Sharma V, Barat D. The treatment of kala-azar during pregnancy. Natl. Med. J. India. 1993;6:263–5.

64. Giri O. Amphotericin B therapy in kala azar.pdf. J. Indian Med. Assoc. 1993;91:91–3.

65. Gradoni L, Gaeta GB, Pellizzer G, Maisto A, Scalone A. Mediterranean visceral leishmaniasis in pregnancy. Scand. J. Infect. Dis. 1994;26:627–9.

66. Jeronimo SMB, Oliveira RM, Mackay S, Costa RM, Sweet J, Eliana T, et al. An urban outbreak of visceral leishmaniasis in Natal, BRazil. Trans. R. Soc. Trop. Med. Hyg. 1994;88:386–8.

67. Sharma R, Bahl L, Goel A, Upadhaya A, Kaushik S, Sharma R, et al. Congenital kala-azar: a case report. J Commun Dis. 1996;Mar:59–61.

68. Thakur C, Kumar P, Kumar N, Singh G, Singh A, Narain S. A randomised comparision of classifical mode of administration of amphotericin B with its newer modes of administration in kala-azar. J. Assoc. Physicians India. 1998;46:779–83.

69. Thakur CP, Singh RK, Hassan SM, Kumar R, Narain S, Kumar A. Amphotericin B deoxycholate treatment of visceral leishmaniasis with newer modes of administration and precautions: A study of 938 cases. Trans. R. Soc. Trop. Med. Hyg. 1999;93:319–23.

70. Meinecke CK, Schottelius J, Oskam L, Fleischer B. Congenital Transmission of Visceral Leishmaniasis (Kala Azar) From an Asymptomatic Mother to Her Child. Pediatrics. 1999;104:e65–e65.

71. Vianna VL, Takiya CM, de Brito-Gitirana L. Histopathologic analysis of hamster hepatocytes submitted to experimental infection with Leishmania donovani. Parasitol. Res. 2002;88:829–36.

72. Dereure J, Duong Thanh H, Lavabre-Bertrand T, Cartron G, Bastides F, Richard-Lenoble D, et al. Visceral leishmaniasis. Persistence of parasites in lymph nodes after clinical cure. J. Infect. 2003;47:77–81.

73. Caldas AJM, Costa JML, Gama MEA, Ramos EAG, Barral A. Visceral leishmaniasis in pregnancy: A case report. Acta Trop. 2003;88:39–43.

74. Pagliano P, Rossi M, Rescigno C, Altieri S, Coppola MG, Gramiccia M, et al. Mediterranean visceral leishmaniasis in HIV-negative adults: A retrospective analysis of 64 consecutive cases (1995-2001). J. Antimicrob. Chemother. 2003;52:264–8.

75. Kumar P V., Daneshbod Y, Sadeghipoor A. Leishmania in the glomerulus. Arch. Pathol. Lab. Med. 2004;128:935–6.

76. Figueiró Filho EA, Uehara SNO, Senefonte FR de A, Lopes AHA, Duarte G, El Beitune P. Leishmaniose visceral e gestação: relato de caso. Rev. Bras. Ginecol. e Obs. 2005;27:92–7.

77. Boehme C, Hain U, Novosel A, Eichenlaub S, Fleischmann E, Loscher T. Congenital visceral leishmaniasis. Emerg. Infect. Dis. 2006;12:359–3.

78. Mueller M, Ritmeijer K, Balasegaram M, Koummuki Y, Santana MR, Davidson R. Unresponsiveness to AmBisome in some Sudanese patients with kala-azar. Trans. R. Soc. Trop. Med. Hyg. 2007;101:19–24.

79. Topno RK, Pandey K, Das VNR, Kumar N, Bimal S, Verma RB, et al. Visceral leishmaniasis in pregnancy — the role of amphotericin B. Ann. Trop. Med. Parasitol. 2008;102:267–70.

80. Lorenzi A, Williams C, Griffiths I. Visceral leishmaniasis mimicking disease activity in mixed connective tissue disease. Rheumatology. 2008;47:737–8.

81. Sinha PK, Roddy P, Palma PP, Kociejowski A, Lima MA, Das VNR, et al. Effectiveness and safety of liposomal amphotericin b for visceral leishmaniasis under routine program conditions in Bihar, India. Am. J. Trop. Med. Hyg. 2010;83:357–64.

82. Haque MA, Ekram ARMS, Sharmin LS, Belaluddin M, Salam MA. Congenital visceral leishmaniasis. Pakistan J. Med. Sci. 2010;26:485–7.

83. Ritmeijer K, Ter Horst R, Chane S, Aderie EM, Piening T, Collin SM, et al. Limited effectiveness of high-dose liposomal amphotericin B (AmBisome) for treatment of visceral leishmaniasis in an ethiopian population with high HIV prevalence. Clin. Infect. Dis. 2011;53.

84. Pilaca A, Delia Z, Pepa A, Puca E, Kraja D. Vertical Transmission of the Visceral Leishmaniasis: A Case Report. US-China Med. Sci. 2011;8:642–5.

85. Damodaran S, Erumbala G, Abraham D, Nirmal S. Incidence of leishmaniasis in a district general hospital. Arch. Dis. Child. 2012;97:A253.

86. Mescouto-Borges MRM, Maués É, Costa DL, da Silva Pranchevicius MC, Romero GAS Congenitally transmitted visceral leishmaniasis: Report of two Brazilian human cases. Brazilian J. Infect. Dis. 2013;17:263–6.

87. Milosevic S, Bogavac M, Malenkovic G, Fabri M, Ruzic M, Dugandzija T. Visceral leishmaniasis as a cause of postpartum pyrexia - Case report. Cent. Eur. J. Med. 2013;8:149– 52.

88. Salih NAW, van Griensven J, Chappuis F, Antierens A, Mumina A, Hammam O, et al. Liposomal amphotericin B for complicated visceral leishmaniasis (kala-azar) in eastern Sudan: How effective is treatment for this neglected disease? Trop. Med. Int. Heal. 2014;19:146–52.

89. Burza S, Sinha PK, Mahajan R, Lima MA, Mitra G, Verma N, et al. Five-Year Field Results and Long-Term Effectiveness of 20 mg/kg Liposomal Amphotericin B (Ambisome) for Visceral Leishmaniasis in Bihar, India. PLoS Negl. Trop. Dis. 2014;8:46.

90. Bode SFN, Bogdan C, Beutel K, Behnisch W, Greiner J, Henning S, et al. Hemophagocytic lymphohistiocytosis in imported pediatric visceral leishmaniasis in a nonendemic area. J. Pediatr. 2014;165:147–153.e1.

91. Chiverto Llamazares Y, Cabezas Lopez E, Castro Sanchez M, Iglesias Goy E. Visceral leishmaniasis as a diagnosis of puerperal fever of unknown origin. [Spanish]. Progresos Obstet. y Ginecol. 2014;57:247–50.

92. Rahman KM, Olsen A, Harley D, Butler CD, Mondal D, Luby SP, et al. Kala-azar in Pregnancy in Mymensingh, Bangladesh: A Social Autopsy. PLoS Negl. Trop. Dis. 2014;8:e2710.

93. Colomba C, Adamoli L, Trizzino M, Siracusa L, Bonura S, Tolomeo M, et al. A case of visceral leishmaniasis and pulmonary tuberculosis in a post-partum woman. Int. J. Infect. Dis. International Society for Infectious Diseases; 2015;33:e5–6.

94. Pawar S, Ragesh R, Nischal N, Sharma S, Panda PK, Sharma SK. Unique triad of ‘pregnancy, kala azar and hemophagocytic lymphohistiocytic syndrome from a non-endemic region.’ J. Assoc. Physicians India. 2015;63:65–8.

95. Kumar R, Kumari S, Prakash J, Kumar R. Atypical presentations of visceral leishmaniasis: A case series and review of literature. Trop. J. Med. Res. 2015;18:109–12.

96. Silva Jde A, Araujo Ide M, Pavanetti LC, Okamoto LS, Dias M. [Visceral leishmaniasis and pregnancy in renal transplanted patient: case report]. J. Bras. Nefrol. 2015;37:268–70.

97. Panagopoulos P, Mitsopoulos V, Papadopoulos A, Theodorou S, Christodoulaki C, Aloupogiannis K, et al. Visceral leishmaniasis during pregnancy: A rare case report from Greece. PLoS Negl. Trop. Dis. 2017;11:e0005134.

98. Adam GK, Omar SM, Ahmed MAA, Abdallah TM, Ali AAA. Cross-sectional study of the case–fatality rate among patients with visceral leishmaniasis infections during pregnancy in Sudan. Int. J. Gynecol. Obstet. 2018;140:119–20.

99. Goyal V, Mahajan R, Pandey K, Singh SN, Singh RS, Strub-Wourgaft N, et al. Field safety and effectiveness of new visceral leishmaniasis treatment regimens within public health facilities in Bihar, India. PLoS Negl. Trop. Dis. 2018;12:e0006830.

100. Russo A, Alt F, Neu MA, Eder S, Wingerter A, Malki KE, et al. Hemophagocytic lymphohistiocytosis in early infancy-pitfall of differentiation between hereditary and infectious reasons. Blood. Conf. 60th Annu. Meet. Am. Soc. Hematol. ASH. 2018;132.

101. Cunha FT, Lopes IC, Oliveira FCS, Queiroz IT. Visceral leishmaniasis in pregnant women from Rio Grande do Norte, Brazil: A case report and literature review. Rev. Soc. Bras. Med. Trop. 2019;52:10–2.

102. Parise ÉV, Maia FSC, Gomes NSG, Silva ACP da. Óbito por leishmaniose visceral em puérpera no município de Palmas, Tocantins, Brasil. J. Heal. Biol. Sci. 2019;7:312–9.

103. Napier LE. Kala-Azar. Princ. Pract. Trop. Med. New York, USA: The Macmillan Company; 1946. p. 141.

104. Napier LE, Das Gupta CR. Indian Kala-Azar in a newly-born child. Ind. Med. Gaz. 1928;April:199–200.

105. Ahluwalia IB, Bern C, Wagatsuma Y, Costa C, Chowdhury R, Ali M, et al. Visceral Leishmaniasis: Consequences to Women in a Bangladeshi Community. J. Women’s Heal. 2004;13:360–4.

106. Office of the director of Census. Census of India 2011 [Internet]. 2011 [cited 2019 Sep 19]. Available from: http://censusindia.gov.in/2011-prov-results/data_files/bihar/Provisional_Population_Totals_2011-Bihar.pdf

107. UNESCO. Ethiopia [Internet]. 2017 [cited 2019 Sep 19]. Available from: http://uis.unesco.org/country/ET

108. WHO TDR. Central registry for epidemiological surveillance of drug safety in pregnancy [Internet]. WHO. 2019 [cited 2019 Sep 19]. Available from: https://www.who.int/tdr/research/tb_hiv/drug-safety-pregnancy/en/

109. Infectious Diseases Data Observatory. Visceral Leishmaniasis: Contributing data [Internet]. [cited 2021 Feb 12]. Available from: https://www.iddo.org/vl/data-sharing/contributing-data

110. Bhattacharya SK, Sinha PK, Sundar S, Thakur CP, Jha TK, Pandey K, et al. Phase 4 Trial of Miltefosine for the Treatment of Indian Visceral Leishmaniasis. J. Infect. Dis. 2007;196:591– 8.

111. Mondal D, Alvar J, Hasnain MG, Hossain MS, Ghosh D, Huda MM, et al. Efficacy and safety of single-dose liposomal amphotericin B for visceral leishmaniasis in a rural public hospital in Bangladesh: A feasibility study. Lancet Glob. Heal. 2014;2:e51–7.

112. Jamil KM, Haque R, Rahman R, Faiz MA, Bhuiyan ATMRH, Kumar A, et al. Effectiveness Study of Paromomycin IM Injection (PMIM) for the Treatment of Visceral Leishmaniasis (VL) in Bangladesh. PLoS Negl. Trop. Dis. 2015;9:1–11.

113. Pandey K, Ravidas V, Siddiqui NA, Sinha SK, Verma RB, Singh TP, et al. Pharmacovigilance of miltefosine in treatment of visceral leishmaniasis in endemic areas of Bihar, India. Am. J. Trop. Med. Hyg. 2016;95:1100–5.

114. Saito M, Gilder ME, Nosten F, Guérin PJ, McGready R. Methodology of assessment and reporting of safety in anti-malarial treatment efficacy studies of uncomplicated falciparum malaria in pregnancy: A systematic literature review. Malar. J. 2017;16:1–10.

