## Supplementary material for "Visceral Leishmaniasis in pregnancy and vertical transmission: A systematic literature review on the therapeutic orphans": S2 Text

**Supplemental file 2:**

**Visceral Leishmaniasis in pregnancy and vertical transmission: A systematic review of the literature on the therapeutic orphans**

Prabin Dahal^1,2*^, Sauman Singh-Phulgenda^1,2^, Brittany J Maguire^1,2^, Eli Harriss^3^, Koert Ritmeijer^4^, Fabiana Alves^5^, Philippe J Guerin^1,2^, Piero L Olliaro^2^

^1^Infectious Diseases Data Observatory (IDDO), Oxford, UK

^2^Centre for Tropical Medicine and Global Health, Nuffield Department of Clinical Medicine,

University of Oxford, Oxford, UK

^3^The Knowledge Centre, Bodleian Health Care Libraries, University of Oxford, Oxford, UK

^4^Médecins Sans Frontières, Amsterdam, Netherlands

^5^Drugs for Neglected Diseases initiative, Geneva, Switzerland

**Section 1: Search strategies**

Date: Search strategies run on 26/03/2020 at the Bodleian Health Care Libraries, University of Oxford.

### Search Strategies

**Database: Medline (Ovid MEDLINE® Epub Ahead of Print, In-Process & Other Non-Indexed Citations, Ovid MEDLINE® Daily and Ovid MEDLINE®) 1946 to present**

Search Strategy:

--------------------------------------------------------------------------------

1	Infectious Disease Transmission, Vertical/ (15643)

2	(vertical* adj4 transmi*).tw. (8130)

3	(perinatal* adj4 (infect* or Acquir* or transmi* or expos*)).tw. (8874)

4	(peri-natal* adj4 (infect* or Acquir* or transmi* or expos*)).tw. (15)

5	(Fetomaternal* adj4 (infect* or Acquir* or transmi* or expos*)).tw. (27)

6	("maternal fetal" adj4 (infect* or Acquir* or transmi* or expos*)).tw. (555)

7	(maternofetal adj4 (infect* or Acquir* or transmi* or expos*)).tw. (129)

8	("mother* to child*" adj4 (infect* or Acquir* or transmi* or expos*)).tw. (6401)

9	("mother* to infant*" adj4 (infect* or Acquir* or transmi* or expos*)).tw. (1094)

10	(intrauterine* adj4 (infect* or Acquir* or transmi* or expos*)).tw. (4830)

11	(intra-uterine* adj4 (infect* or Acquir* or transmi* or expos*)).tw. (295)

12	(utero adj4 (infect* or Acquir* or transmi* or expos*)).tw. (8528)

13	1 or 2 or 3 or 4 or 5 or 6 or 7 or 8 or 9 or 10 or 11 or 12 (42542)

14	exp Pregnancy/ or exp Pregnant Women/ (884794)

15	(pregnan* or gravid*).ti,ab. (502424)

16	matern*.ti,ab. (270558)

17	14 or 15 or 16 (1096954)

18	13 or 17 (1110427)

19	Leishmaniasis, Visceral/ (9748)

20	"black fever".ti,ab. (15)

21	"kala azar".ti,ab. (2670)

22	"visceral leishmaniasis".ti,ab. (8202)

23	19 or 20 or 21 or 22 (12280)

24	18 and 23 (145)

**Database: Embase 1974 to present**

Search Strategy:

--------------------------------------------------------------------------------

1	vertical transmission/ (14592)

2	(vertical* adj4 transmi*).tw. (9699)

3	(perinatal* adj4 (infect* or Acquir* or transmi* or expos*)).tw. (11165)

4	(peri-natal* adj4 (infect* or Acquir* or transmi* or expos*)).tw. (28)

5	(Fetomaternal* adj4 (infect* or Acquir* or transmi* or expos*)).tw. (35)

6	("maternal fetal" adj4 (infect* or Acquir* or transmi* or expos*)).tw. (666)

7	(maternofetal adj4 (infect* or Acquir* or transmi* or expos*)).tw. (169)

8	("mother* to child*" adj4 (infect* or Acquir* or transmi* or expos*)).tw. (7927)

9	("mother* to infant*" adj4 (infect* or Acquir* or transmi* or expos*)).tw. (1299)

10	(intrauterine* adj4 (infect* or Acquir* or transmi* or expos*)).tw. (6334)

11	(intra-uterine* adj4 (infect* or Acquir* or transmi* or expos*)).tw. (371)

12	(utero adj4 (infect* or Acquir* or transmi* or expos*)).tw. (11153)

13	1 or 2 or 3 or 4 or 5 or 6 or 7 or 8 or 9 or 10 or 11 or 12 (50331)

14	exp pregnancy/ or pregnant woman/ (689555)

15	(pregnan* or gravid*).ti,ab. (628232)

16	matern*.ti,ab. (341175)

17	14 or 15 or 16 (1037206)

18	13 or 17 (1058932)

19	exp visceral leishmaniasis/ (9608)

20	"black fever".ti,ab. (17)

21	"kala azar".ti,ab. (2218)

22	"visceral leishmaniasis".ti,ab. (9299)

23	19 or 20 or 21 or 22 (12302)

24	18 and 23 (163)

Cochrane Database of Systematic Reviews

Issue 3 of 12, March 2020

Cochrane Central Register of Controlled Trials

Issue 3 of 12, March 2020

#1	MeSH descriptor: [Infectious Disease Transmission, Vertical] explode all trees	525

#2	vertical* near/4 transmi*	792

#3	perinatal* near/4 (infect* or Acquir* or transmi* or expos*)	517

#4	peri-natal* near/4 (infect* or Acquir* or transmi* or expos*)	0

#5	Fetomaternal* near/4 (infect* or Acquir* or transmi* or expos*)	6

#6	("maternal fetal" near/4 (infect* or Acquir* or transmi* or expos*))	45

#7	maternofetal near/4 (infect* or Acquir* or transmi* or expos*)	8

#8	("mother* to child*" near/4 (infect* or Acquir* or transmi* or expos*))	663

#9	("mother* to infant*" near/4 (infect* or Acquir* or transmi* or expos*))	49

#10	(intrauterine* near/4 (infect* or Acquir* or transmi* or expos*))	316

#11	(intra-uterine* near/4 (infect* or Acquir* or transmi* or expos*))	13

#12	utero near/4 (infect* or Acquir* or transmi* or expos*)	301

#13	#1 or #2 or #3 or #4 or #5 or #6 or #7 or #8 or #9 or #10 or #11 or #12	2071

#14	MeSH descriptor: [Pregnancy] explode all trees	7602

#15	MeSH descriptor: [Pregnant Women] explode all trees	242

#16	pregnan* or gravid* or matern*	71901

#17	#14 or #15 or #16	72090

#18	#13 or #17	72564

#19	MeSH descriptor: [Leishmaniasis, Visceral] explode all trees	37

#20	"black fever"	1

#21	"kala azar"	118

#22	"visceral leishmaniasis"	241

#23	#19 or #20 or #21 or #22	282

#24	#18 and #23	14

World Health Organization Global Index Medicus: LILACS (Americas); IMSEAR (South-East Asia); IMEMR (Eastern Mediterranean); WPRIM (Western Pacific)

https://www.globalindexmedicus.net/

Title, abstract, subject: ("black fever" OR "kala azar" OR "visceral leishmaniasis") AND (pregnan* OR gravid* OR matern* OR "vertical* transmi*" OR perinatal* OR peri-natal* OR fetomatern* OR mother* OR intrauterine OR intra-uterine OR utero)

ClinicalTrials.gov Advanced Search <https://clinicaltrials.gov/ct2/search/advanced>?

Condition or disease: visceral leishmaniasis

Other terms: pregnant

=2

Condition or disease: visceral leishmaniasis

Other terms: pregnancy

=1

Condition or disease: visceral leishmaniasis

Other terms: maternal

=0

Condition or disease: visceral leishmaniasis

Other terms: maternity

=1

Condition or disease: visceral leishmaniasis

Other terms: gravid

=2

Condition or disease: visceral leishmaniasis

Other terms: vertical transmission

=0

Condition or disease: visceral leishmaniasis

Other terms: perinatal

=0

Condition or disease: visceral leishmaniasis

Other terms: fetomaternal

=0

Condition or disease: visceral leishmaniasis

Other terms: mother

=0

Condition or disease: visceral leishmaniasis

Other terms: peri-natal

=0

Condition or disease: visceral leishmaniasis

Other terms: intrauterine

=0

Condition or disease: visceral leishmaniasis

Other terms: intra-uterine

=0

Condition or disease: visceral leishmaniasis

Other terms: utero

=0

Condition or disease: visceral leishmaniasis

Other terms: mothers

=0

WHO International Clinical Trials Registry Platform <https://www.who.int/ictrp/en/>

#### Important information related to the COVID-19 outbreak!

Due to heavy traffic generated by the COVID-19 outbreak, the ICTRP Search Portal is not accessible from outside WHO temporarily. Please subscribe to the ICTRP listserv if you wish to be notified when the search portal is working again. Information on how to subscribe can be found on the same page below.

### Search Results

| Ovid Medline | 145 |
| --- | --- |
| Ovid Embase | 163 |
| Cochrane Database of Systematic Reviews and Cochrane CENTRAL | 14 |
| Global Index Medicus | 67 |
| Clinicaltrials.gov | 6 |
| TOTAL | 395 |
| Total after deduplication | 272 |

Manual search:

ISRCTN registry <https://www.isrctn.com/search?q=visceral+leishmaniasis&searchType=advanced-search>

Text search: visceral leishmaniasis

**Section 2: Study inclusion and exclusion criteria**

**Table 2: Selection criteria for published study reports on visceral Leishmaniasis**

| **Inclusion criteria** | **Exclusion criteria** |
| --- | --- |
| All studies involving Visceral Leishmaniasis in pregnant women or cases of congenital transmission | Animal studies |
| All antileishmanial formulations (regardless of whether or not treatment was administered) | Non-clinical studies (descriptions of laboratory methods, modelling studies, economic evaluations) |
|  | Publications and trials describing studies on cutaneous leishmaniasis, post kala-azar dermal leishmaniasis (PKDL), canine VL, vector control, nets, prevalence estimation, diagnostic tests, vaccines or prophylaxis |
